# Detection and clearance of type-specific and phylogenetically related genital human papillomavirus infections in young women in new heterosexual relationships

**DOI:** 10.1101/2023.02.24.23286387

**Authors:** Andrew W. Arthur, Mariam El-Zein, Ann N. Burchell, Pierre-Paul Tellier, Francois Coutlée, Eduardo L. Franco

## Abstract

**Background:** Understanding the natural history of human papillomavirus (HPV) infections is essential to effective cervical cancer prevention planning. We examined these outcomes in-depth among young women.

**Methods:** The HPV Infection and Transmission among Couples through Heterosexual Activity (HITCH) study is a prospective cohort of 501 college-age women who recently initiated a heterosexual relationship. We tested vaginal samples collected at six clinical visits over 24 months for 36 HPV types. Using rates and Kaplan-Meier analysis, we estimated time-to-event statistics with 95% confidence intervals (CIs) for detection of incident infections and liberal clearance of incident and present-at-baseline infections (separately). We conducted analyses at the woman- and HPV-levels, with HPV types grouped by phylogenetic relatedness.

**Results:** By 24 months, we detected incident infections in 40.4%, CI:33.4-48.4 of women. Incident subgenus 1 (43.4, CI:33.6-56.4), 2 (47.1, CI:39.9-55.5) and 3 (46.6, CI:37.7-57.7) infections cleared at similar rates per 1000 infection-months. We observed similar homogeny in HPV-level clearance rates among present-at-baseline infections.

**Conclusions:** Our woman-level analyses of infection detection and clearance agreed with similar studies. However, our HPV-level analyses did not clearly indicate that high oncogenic risk subgenus 2 infections take longer to clear than their low oncogenic risk and commensal subgenera 1 and 3 counterparts.

## INTRODUCTION

In 2020, cervical cancer accounted for 3.1% of the global cancer burden (604,127 cases) [1]. Persistent genital infection with oncogenic types of human papillomavirus (HPV) is a necessary cause of the majority of cervical precancerous lesions and cancers [2-6]. While most infections in young women are transient, a minority persist [7], and a sizeable proportion of “incident” infections in older women are reactivations of previously-undetectable infections acquired earlier in life [8].

Cervical cancer is a highly preventable disease. The nonavalent HPV vaccine prevents infection with HPV types found in 89.5% of invasive cervical cancers [9], and molecular HPV testing is an efficacious screening strategy [10]. Studies of HPV infection natural history in young women [11-23] have provided parameters for models and algorithms that inform primary and secondary cervical cancer prevention strategies.

In these studies, the woman was the unit of observation (i.e., analyses were conducted at the woman-level). Woman-level analyses incorporate the detection or clearance of multiple HPV infections into composite outcomes, which can obscure detections and clearances of multiple individual infections in the same woman. HPV-level analyses (where the HPV infection is the unit of observation), have a stronger biological rationale for characterising infection natural history because they treat the detection and clearance of each unique infection as a separate HPV type-specific event.

We described vaginal HPV infection prevalence, incidence (i.e., time to detection), and persistence (i.e., time to clearance) with respect to individual HPV types, as well as types grouped by subgeneric classifications using woman-level and HPV-level paradigms. We based our analyses on a cohort of women who had recently initiated a new sexual relationship with a male partner in the HPV Infection and Transmission among Couples through Heterosexual activity (HITCH) study.

## METHODS

### Study Design and Procedures

We used data from female participants of the HITCH prospective cohort study. Study details have been published elsewhere [24]. Briefly, we recruited female university and college students (aged 18-24) who began a sexual relationship with a male partner ≤ 6 months prior. Enrolment occurred between 2005 and 2011 in Montréal, Canada, at university-run health clinics. At baseline, 4-, 8-, 12-, 18- and 24-months post-enrollment, women provided a vaginal sample and completed sociodemographic/sexual behavioural questionnaires. Women were asked to refrain from sexual activity for 24 hours before each visit. Based on nurse instructions, participants self-collected vaginal samples using a polyester swab. The diagnostic accuracy of self-sampling has been demonstrated [25-27]. We used the Linear Array genotyping assay (Roche Molecular Systems, CA) to detect (individually) 36 HPV types [28], validating samples via β-globin DNA coamplification. After the 2006 licensing of the HPV vaccine, women were asked how many doses they had received. The HITCH study was approved by the Institutional Review Boards of McGill and Concordia Universities as well as the Centre Hospitalier de l’Université de Montreal; participants provided written informed consent.

### Taxonomic Groups

We described the natural history of individual HPV types separately. We did the same for three groups of phylogenetically related HPV types, as defined by subgenera of the Alphapapillomavirus genus. This taxonomic scheme clusters HPV types according to tissue tropism and oncogenic risk, based on empirical evidence and differential mucosotropic type distributions. Subgenus 1 includes low oncogenic risk HPVs 6, 11, 40, 42, 44, and 54; subgenus includes high oncogenic risk HPVs 16, 18, 26, 31, 33, 35, 39, 45, 51, 52, 53, 56, 58, 59, 66, 67, 68, 69, 70, 73 and 82; and subgenus 3 includes commensal HPVs 61, 62, 71, 72, 81, 83, 84 and 89 [29-31]. Analyses designated “Any HPV” or “All HPV”, incorporate all 36 types.

### Analytical Frameworks

We performed analyses on the woman-level (see Addenda 1 and 2) and the HPV-level (see Addendum 3). We analysed the outcome “single detection” of incident infections (Figure 1A). Woman-level detection occurred the first time a woman tested positive for the HPV type(s) of interest, whereas HPV-level detection occurred each first time a woman tested positive for a distinct type of interest. To restrict to incident detections, we excluded women positive for the type(s) of interest at baseline from woman-level analyses, and types of interest women were positive for at baseline from HPV-level analyses.

**Figure 1A.**
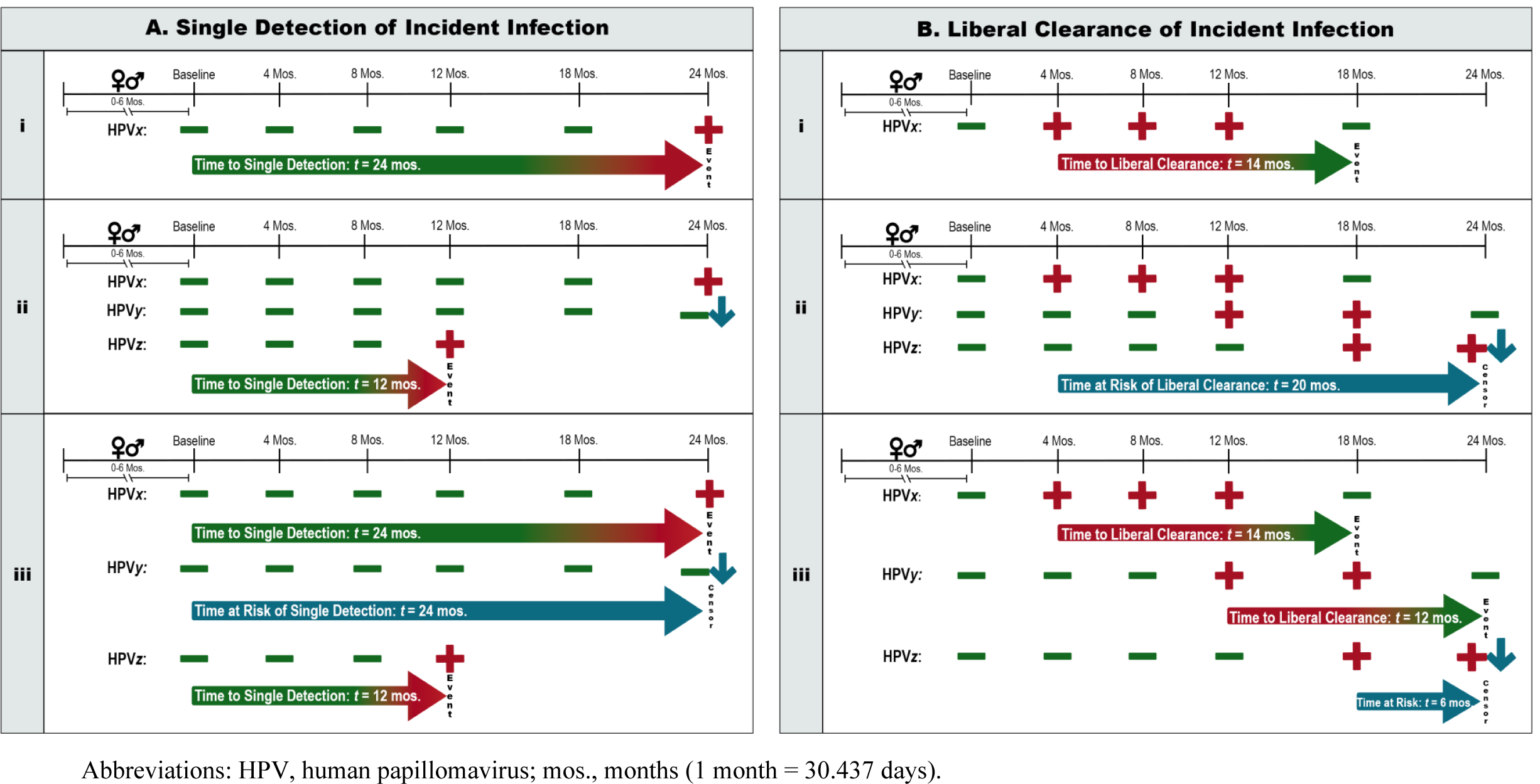
Analytical framework for single detection of incident infection. **i.** Woman-level analysis for a given HPV type: Woman A must be negative for HPV*x* at baseline. She has a single detection at 24 months, when she tests positive for HPV*x*. **ii.** Woman-level analysis for grouped HPV types *x, y*, and *z*: Woman A must be negative for all three types at baseline. She has a single detection at 12 months, the first time she tests positive for one (or more) types following a visit where she was negative for all three types. **iii.** HPV-level analysis for grouped HPV types *x, y*, and *z*: HPV types must be absent at baseline. There will be a single detection each first time woman A tests positive for any type following a visit where she was negative for the same type. HPV*x* and HPV*z* have single detections at 24- and 12-months, respectively; HPV*y* is right-censored (24 months). **Figure 1B**. Analytical framework for liberal clearance of incident infection. **i.** Woman-level analysis for a given HPV type: Woman B must be negative for HPV*x* at baseline. She later tests positive for HPV*x*, then has a liberal clearance after 14 months have elapsed, when she tests negative for HPV*x*. **ii.** Woman-level analysis for grouped HPV types *x, y*, and *z*: Woman B must be negative for all three types at baseline. After testing positive for HPV*x*, she never clears all three asynchronously-detected infections at a single visit. She is right-censored (24 months). **iii.** HPV-level analysis for grouped HPV types *x, y*, and *z*: HPV types must be absent at baseline, then detected later in the study. There will be a liberal clearance each first time woman B tests negative for any type following a visit where she was positive for the same type. HPV*x* and HPV*y* have liberal clearances 14- and 12-months post-detection, respectively; HPV*z* is right-censored 6 months after detection. Symbol Legend: 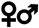: Debut of woman’s sexual relationship with a male partner occurred 0-6 months pre-baseline. 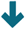: Data are right-censored. 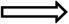: Time to discrete event of interest. Gradient arrows indicate that biological infection/clearance occurred at an unknown time before detection/clearance at the arrowhead. Solid blue arrow counts time at risk contributed before censorship at the arrowhead.

We then analyzed the outcome “liberal clearance.” Woman-level clearance occurred the first time a woman tested negative for all type(s) of interest after testing positive for one (or more) types of interest, whereas HPV-level clearance occurred each first time a woman tested negative for a distinct type of interest following a positive test for the same type. We analysed liberal clearance in the context of infections present at baseline (Figure S1) and incident infections (Figure 1B) separately. To restrict to infections present at baseline, we included women who were positive for the type(s) of interest at baseline from woman-level analyses, and types of interest women were positive for at baseline from HPV-level analyses. To restrict to incident infections, we included women who were initially negative, then later positive for the type(s) of interest in woman-level analyses, and types of interest for which women were initially negative, then later positive in HPV-level analyses.

We repeated all analyses substituting more stringent outcome definitions: double detection and conservative clearance. These outcomes were akin to single detection and liberal clearance (respectively) on the woman- and HPV-levels, however, the defining positive/negative results occurred at two consecutive visits. We measured times to these more-stringent events based on the first of the two positive/negative visits.

### Statistical Analyses

For all HPV-level analyses, we accounted for potential intraparticipant correlation arising from repeated measurement of multiple HPV types for the same woman. We calculated, by visit, HPV prevalence with 95% exact confidence intervals (CIs). For HPV-level analyses, we accounted for intra-woman clustering using a degrees of freedom-adjusted effective sample size [32].

We estimated Kaplan-Meier (KM) product-limit rates at 6, 12, and 24 months to describe cumulative detection and percent of infections uncleared. Using the log-log approach, we assigned 95% CIs to woman-level estimates. For HPV-level estimates, we assigned pointwise percentile-based woman-clustered bootstrap 95% CIs [33, 34].

We estimated the rate of detection/clearance per 1000 woman-months for woman-level analyses, and per 1000 infection-months for HPV-level analyses. 95% CIs around rates were estimated via quadratic approximation of Poisson log-likelihood for woman-level analyses. We extracted 95% CIs from a leave-one-woman-cluster-out jackknife procedure for HPV-level analyses.

We calculated all mean and median times to detection/clearance including censored observations (i.e., actuarial measures of central tendency), then excluding censored observations (i.e., outcome-conditional measures of central tendency). We estimated mean times to detection/clearance (restricted to longest follow-up) with parametric 95% CIs for woman-level analyses, and percentile-based woman-clustered bootstrap 95% CIs for HPV-level analyses. Based on the log-log survival function CIs, we assigned 95% CIs to the woman-level median, and based on the woman-clustered bootstrap survival function CIs, we assigned 95% CIs to the HPV-level median.

We performed sensitivity analyses to approximate the extent to which right censoring caused actuarial mean times to detection/clearance to be underestimated. We calculated a separate mean time to each event based on the area under a KM survival function with a fitted exponential decay to *S*(*t*) = 0 (hereafter exponentially-extended mean). For woman-level analyses, we assigned percentile-based bootstrap 95% CIs; for HPV-level analyses, bootstraps were resampled by woman-clusters. Statistical analyses were conducted using Stata SE 17.0 (StataCorp LLC., TX).

## RESULTS

Of 502 women enrolled, 453 provided two or more valid vaginal samples; 48 had only one valid sample and were included in prevalence estimates but not survival analyses. For six women missing a valid vaginal sample at baseline, we treated samples provided at visit 2 as baseline samples. Loss-to-follow-up by visit 6 was 43.7% (Figure S2). Among the 453 women included in survival analyses, median follow-up was 26.4 (quartiles 1-3: 19.5-31.7) months. Most women identified with the following ethnicities: English Canadian (34.0%), French Canadian (27.3%), Italian (4.6%), Latin American (4.9%), and Multiple or Mixed Ethnicities (5.1%). At baseline, the mean age was 20.7 years (standard deviation: 1.8). Women reported an average weekly vaginal intercourse frequency of 4.6 and, on average, 6.4 previous heterosexual relationships involving vaginal sex. Among 93 women who reported having received the HPV vaccine, the mean number of doses was 2.6.

As shown in Table 1, the three most prevalent HPV types at baseline were HPV16 (16.8%, CI:13.6-20.3), HPV89 (10.6%, CI:8.0-13.6), and HPV51 (10.2%, CI:7.7-13.2). Across follow-up, HPV16 (10.7-16.8%) and HPV89 (9.5-10.6%) remained among the three most-prevalent types. At baseline, the prevalence of any HPV infection was 57.1%, CI:52.6-61.5 at the woman-level (n_w_=501), and 4.3%, CI:3.9-4.8 at the HPV-level (n_HPV_=18,036).

**Table 1.** Prevalence at each visit [n_positive_, % (95% CI)] of individual HPV types, grouped types at the woman-level, and grouped types at the HPV level per visit, by subgenus. Sample size at each visit is reported for woman- (n_w_) and HPV- (n_HPV_) level analyses.

|  | Visit 1 | Visit 2 | Visit 3 | Visit 4 | Visit 5 | Visit 6 |
| --- | --- | --- | --- | --- | --- | --- |
| | $n_w=501$, $n_{\text{HPV}}=18,036$ | $n_w=451$, $n_{\text{HPV}}=16,236$ | $n_w=412$, $n_{\text{HPV}}=14,832$ | $n_w=374$, $n_{\text{HPV}}=13,464$ | $n_w=326$, $n_{\text{HPV}}=11,736$ | $n_w=282$, $n_{\text{HPV}}=10,152$ |
| **Subgenus 1** |  |  |  |  |  |  |
| HPV6 | 19, 3.8 (2.3, 5.9) | 17, 3.8 (2.2, 6.0) | 19, 4.6 (2.8, 7.1) | 22, 5.9 (3.7, 8.8) | 21, 6.4 (4.0, 9.7) | 18, 6.4 (3.8, 9.9) |
| HPV11 | 3, 0.6 (0.1, 1.7) | 0, 0.0 (0.0, 0.8)<sup>a</sup> | 0, 0.0 (0.0, 0.9)<sup>a</sup> | 1, 0.3 (0.0, 1.5) | 2, 0.6 (0.1, 2.2) | 1, 0.4 (0.0, 2.0) |
| HPV40 | 12, 2.4 (1.2, 4.2) | 6, 1.3 (0.5, 2.9) | 9, 2.2 (1.0, 4.1) | 12, 3.2 (1.7, 5.5) | 8, 2.5 (1.1, 4.8) | 7, 2.5 (1.0, 5.1) |
| HPV42 | 38, 7.6 (5.4, 10.3) | 43, 9.5 (7.0, 12.6) | 32, 7.8 (5.4, 10.8) | 21, 5.6 (3.5, 8.5) | 24, 7.4 (4.8, 10.8) | 23, 8.2 (5.2, 12.0) |
| HPV44 | 10, 2.0 (1.0, 3.6) | 6, 1.3 (0.5, 2.9) | 5, 1.2 (0.4, 2.8) | 5, 1.3 (0.4, 3.1) | 7, 2.2 (0.9, 4.4) | 5, 1.8 (0.6, 4.1) |
| HPV54 | 30, 6.0 (4.1, 8.4) | 23, 5.1 (3.3, 7.6) | 16, 3.9 (2.2, 6.2) | 18, 4.8 (2.9, 7.5) | 16, 4.9 (2.8, 7.9) | 14, 5.0 (2.7, 8.2) |
| Any Woman-Level | 98, 19.6 (16.2, 23.3) | 80, 17.7 (14.3, 21.6) | 72, 17.5 (13.9, 21.5) | 65, 17.4 (13.7, 21.6) | 65, 19.9 (15.7, 24.7) | 56, 19.9 (15.4, 25.0) |
| Any HPV-Level<sup>b</sup> | 112, 0.6 (0.5, 0.8) | 95, 0.6 (0.5, 0.7) | 81, 0.6 (0.4, 0.7) | 79, 0.6 (0.5, 0.8) | 78, 0.7 (0.5, 0.8) | 68, 0.7 (0.5, 0.9) |
| **Subgenus 2** |  |  |  |  |  |  |
| HPV16 | 84, 16.8 (13.6, 20.3) | 66, 14.6 (11.5, 18.2) | 59, 14.3 (11.1, 18.1) | 40, 10.7 (7.8, 14.3) | 37, 11.4 (8.1, 15.3) | 33, 11.7 (8.2, 16.0) |
| HPV18 | 18, 3.6 (2.1, 5.6) | 13, 2.9 (1.5, 4.9) | 8, 1.9 (0.8, 3.8) | 9, 2.4 (1.1, 4.5) | 11, 3.4 (1.7, 6.0) | 10, 3.6 (1.7, 6.4) |
| HPV26 | 0, 0.0 (0.0, 0.7)<sup>a</sup> | 0, 0.0 (0.0, 0.8)<sup>a</sup> | 0, 0.0 (0.0, 0.9)<sup>a</sup> | 1, 0.3 (0.0, 1.5) | 0, 0.0 (0.0, 1.1)<sup>a</sup> | 0, 0.0 (0.0, 1.3)<sup>a</sup> |
| HPV31 | 24, 4.8 (3.1, 7.0) | 25, 5.5 (3.6, 8.1) | 20, 4.9 (3.0, 7.4) | 15, 4.0 (2.3, 6.5) | 17, 5.2 (3.1, 8.2) | 12, 4.3 (2.2, 7.3) |
| HPV33 | 8, 1.6 (0.7, 3.1) | 2, 0.4 (0.1, 1.6) | 4, 1.0 (0.3, 2.5) | 2, 0.5 (0.1, 1.9) | 0, 0.0 (0.0, 1.1)<sup>a</sup> | 1, 0.4 (0.0, 2.0) |
| HPV34 | 3, 0.6 (0.1, 1.7) | 1, 0.2 (0.0, 1.2) | 0, 0.0 (0.0, 0.9)<sup>a</sup> | 0, 0.0 (0.0, 1.0)<sup>a</sup> | 2, 0.6 (0.1, 2.2) | 0, 0.0 (0.0, 1.3)<sup>a</sup> |
| HPV35 | 4, 0.8 (0.2, 2.0) | 4, 0.9 (0.2, 2.3) | 5, 1.2 (0.4, 2.8) | 5, 1.3 (0.4, 3.1) | 3, 0.9 (0.2, 2.7) | 1, 0.4 (0.0, 2.0) |
| HPV39 | 34, 6.8 (4.8, 9.4) | 32, 7.1 (4.9, 9.9) | 18, 4.4 (2.6, 6.8) | 22, 5.9 (3.7, 8.8) | 15, 4.6 (2.6, 7.5) | 15, 5.3 (3.0, 8.6) |
| HPV45 | 8, 1.6 (0.7, 3.1) | 8, 1.8 (0.8, 3.5) | 3, 0.7 (0.2, 2.1) | 5, 1.3 (0.4, 3.1) | 6, 1.8 (0.7, 4.0) | 8, 2.8 (1.2, 5.5) |
| HPV51 | 51, 10.2 (7.7, 13.2) | 41, 9.1 (6.6, 12.1) | 32, 7.8 (5.4, 10.8) | 20, 5.4 (3.3, 8.1) | 19, 5.8 (3.6, 9.0) | 19, 6.7 (4.1, 10.3) |
| HPV52 | 35, 7.0 (4.9, 9.6) | 28, 6.2 (4.2, 8.9) | 29, 7.0 (4.8, 10.0) | 22, 5.9 (3.7, 8.8) | 15, 4.6 (2.6, 7.5) | 15, 5.3 (3.0, 8.6) |
| HPV53 | 36, 7.2 (5.1, 9.8) | 35, 7.8 (5.5, 10.6) | 35, 8.5 (6.0, 11.6) | 29, 7.8 (5.3, 11.0) | 23, 7.1 (4.5, 10.4) | 16, 5.7 (3.3, 9.1) |
| HPV56 | 25, 5.0 (3.3, 7.3) | 21, 4.7 (2.9, 7.0) | 15, 3.6 (2.1, 5.9) | 13, 3.5 (1.9, 5.9) | 11, 3.4 (1.7, 6.0) | 7, 2.5 (1.0, 5.1) |
| HPV58 | 23, 4.6 (2.9, 6.8) | 20, 4.4 (2.7, 6.8) | 16, 3.9 (2.2, 6.2) | 16, 4.3 (2.5, 6.9) | 13, 4.0 (2.1, 6.7) | 9, 3.2 (1.5, 6.0) |
| HPV59 | 30, 6.0 (4.1, 8.4) | 21, 4.7 (2.9, 7.0) | 16, 3.9 (2.2, 6.2) | 11, 2.9 (1.5, 5.2) | 15, 4.6 (2.6, 7.5) | 11, 3.9 (2.0, 6.9) |
| HPV66 | 31, 6.2 (4.2, 8.7) | 30, 6.7 (4.5, 9.4) | 20, 4.9 (3.0, 7.4) | 20, 5.4 (3.3, 8.1) | 16, 4.9 (2.8, 7.9) | 12, 4.3 (2.2, 7.3) |
| HPV67 | 28, 5.6 (3.8, 8.0) | 22, 4.9 (3.1, 7.3) | 20, 4.9 (3.0, 7.4) | 16, 4.3 (2.5, 6.9) | 14, 4.3 (2.4, 7.1) | 5, 1.8 (0.6, 4.1) |
| HPV68 | 14, 2.8 (1.5, 4.6) | 11, 2.4 (1.2, 4.3) | 13, 3.2 (1.7, 5.3) | 10, 2.7 (1.3, 4.9) | 8, 2.5 (1.1, 4.8) | 7, 2.5 (1.0, 5.1) |
| HPV69 | 0, 0.0 (0.0, 0.7)<sup>a</sup> | 0, 0.0 (0.0, 0.8)<sup>a</sup> | 0, 0.0 (0.0, 0.9)<sup>a</sup> | 0, 0.0 (0.0, 1.0)<sup>a</sup> | 0, 0.0 (0.0, 1.1)<sup>a</sup> | 0, 0.0 (0.0, 1.3)<sup>a</sup> |
| HPV70 | 4, 0.8 (0.2, 2.0) | 3, 0.7 (0.1, 1.9) | 5, 1.2 (0.4, 2.8) | 5, 1.3 (0.4, 3.1) | 5, 1.5 (0.5, 3.5) | 3, 1.1 (0.2, 3.1) |
| HPV73 | 18, 3.6 (2.1, 5.6) | 17, 3.8 (2.2, 6.0) | 19, 4.6 (2.8, 7.1) | 13, 3.5 (1.9, 5.9) | 11, 3.4 (1.7, 6.0) | 11, 3.9 (2.0, 6.9) |
| HPV82 | 14, 2.8 (1.5, 4.6) | 7, 1.6 (0.6, 3.2) | 5, 1.2 (0.4, 2.8) | 6, 1.6 (0.6, 3.5) | 3, 0.9 (0.2, 2.7) | 2, 0.7 (0.1, 2.5) |
| Any Woman-Level | 240, 47.9 (43.5, 52.4) | 207, 45.9 (41.2, 50.6) | 192, 46.6 (41.7, 51.6) | 151, 40.4 (35.4, 45.5) | 139, 42.6 (37.2, 48.2) | 107, 37.9 (32.3, 43.9) |
| Any HPV-Level<sup>b</sup> | 492, 2.7 (2.4, 3.1) | 407, 2.5 (2.2, 2.9) | 342, 2.3 (2.0, 2.7) | 280, 2.1 (1.8, 2.5) | 244, 2.1 (1.8, 2.5) | 197, 1.9 (1.6, 2.3) |
| **Subgenus 3** |  |  |  |  |  |  |
| HPV61 | 12, 2.4 (1.2, 4.2) | 13, 2.9 (1.5, 4.9) | 15, 3.6 (2.1, 5.9) | 13, 3.5 (1.9, 5.9) | 12, 3.7 (1.9, 6.3) | 14, 5.0 (2.7, 8.2) |
| HPV62 | 40, 8.0 (5.8, 10.7) | 43, 9.5 (7.0, 12.6) | 32, 7.8 (5.4, 10.8) | 28, 7.5 (5.0, 10.6) | 24, 7.4 (4.8, 10.8) | 22, 7.8 (5.0, 11.6) |
| HPV71 | 2, 0.4 (0.1, 1.4) | 1, 0.2 (0.0, 1.2) | 0, 0.0 (0.0, 0.9)<sup>a</sup> | 0, 0.0 (0.0, 1.0)<sup>a</sup> | 0, 0.0 (0.0, 1.1)<sup>a</sup> | 0, 0.0 (0.0, 1.3)<sup>a</sup> |
| HPV72 | 2, 0.4 (0.1, 1.4) | 2, 0.4 (0.1, 1.6) | 1, 0.2 (0.0, 1.3) | 1, 0.3 (0.0, 1.5) | 1, 0.3 (0.0, 1.7) | 1, 0.4 (0.0, 2.0) |
| HPV81 | 8, 1.6 (0.7, 3.1) | 7, 1.6 (0.6, 3.2) | 4, 1.0 (0.3, 2.5) | 0, 0.0 (0.0, 1.0)<sup>a</sup> | 0, 0.0 (0.0, 1.1)<sup>a</sup> | 3, 1.1 (0.2, 3.1) |
| HPV83 | 9, 1.8 (0.8, 3.4) | 10, 2.2 (1.1, 4.0) | 6, 1.5 (0.5, 3.1) | 8, 2.1 (0.9, 4.2) | 4, 1.2 (0.3, 3.1) | 3, 1.1 (0.2, 3.1) |
| HPV84 | 48, 9.6 (7.2, 12.5) | 38, 8.4 (6.0, 11.4) | 32, 7.8 (5.4, 10.8) | 19, 5.1 (3.1, 7.8) | 14, 4.3 (2.4, 7.1) | 24, 8.5 (5.5, 12.4) |
| HPV89 | 53, 10.6 (8.0, 13.6) | 45, 10.0 (7.4, 13.1) | 39, 9.5 (6.8, 12.7) | 36, 9.6 (6.8, 13.1) | 26, 8.0 (5.3, 11.5) | 23, 8.2 (5.2, 12.0) |
| Any Woman-Level | 133, 26.6 (22.7, 30.6) | 120, 26.6 (22.6, 30.9) | 99, 24.0 (20.0, 28.5) | 77, 20.6 (16.6, 25.1) | 65, 19.9 (15.7, 24.7) | 72, 25.5 (20.6, 31.0) |
| Any HPV-Level<sup>b</sup> | 174, 1.0 (0.8, 1.1) | 159, 1.0 (0.8, 1.2) | 129, 0.9 (0.7, 1.1) | 105, 0.8 (0.6, 1.0) | 81, 0.7 (0.5, 0.9) | 90, 0.9 (0.7, 1.1) |
| **All 36 Types** |  |  |  |  |  |  |
| Any Woman-Level | 286, 57.1 (52.6, 61.5) | 249, 55.2 (50.5, 59.9) | 227, 55.1 (50.2, 60.0) | 192, 51.3 (46.1, 56.5) | 168, 51.5 (46.0, 57.1) | 141, 50.0 (44.0, 56.0) |
| Any HPV-Level<sup>b</sup> | 778, 4.3 (3.9, 4.8) | 661, 4.1 (3.6, 4.6) | 552, 3.7 (3.3, 4.2) | 464, 3.5 (2.9, 4.0) | 403, 3.4 (2.9, 4.0) | 355, 3.5 (3.0, 4.1) |

Abbreviations: CI, confidence interval; HPV, human papillomavirus.
<sup>a</sup> Type not detected. One-sided 97.5% CI assigned.
<sup>b</sup> Degrees of freedom-adjusted effective sample size used to account for intra-woman clustering.

Table 2 summarizes single detection of incident HPV infections. Among women negative for all HPV types at baseline, cumulative detection of any incident infection was 40.4%, CI:33.4– 48.4 by 24 months. The detection rate for any type was 20.0, CI:16.1-24.9 per 1000 woman-months. Rates per 1000 woman-months were higher for subgenus 2 types (17.0, CI:13.7-20.9) compared to subgenera 1 (11.4, CI:9.3-13.9) and 3 (12.4, CI:10.1-15.1). KM graphs for single detection are displayed in Figure 2 (woman-level) and Figure S3 (HPV-level). Table S1 alongside Figures S4 and S5 reproduce the aforementioned results for double detection analyses.

**Table 2.**
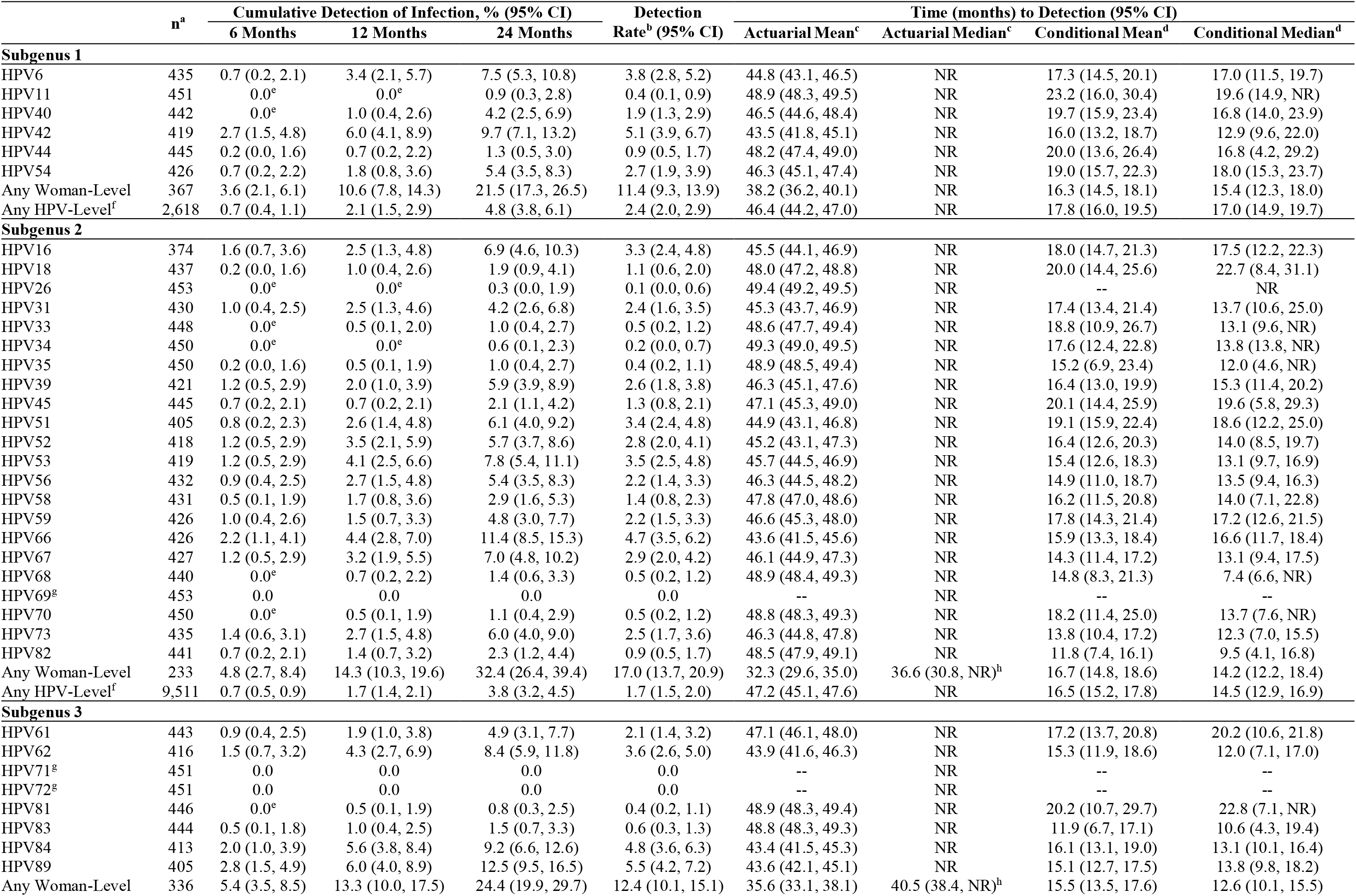
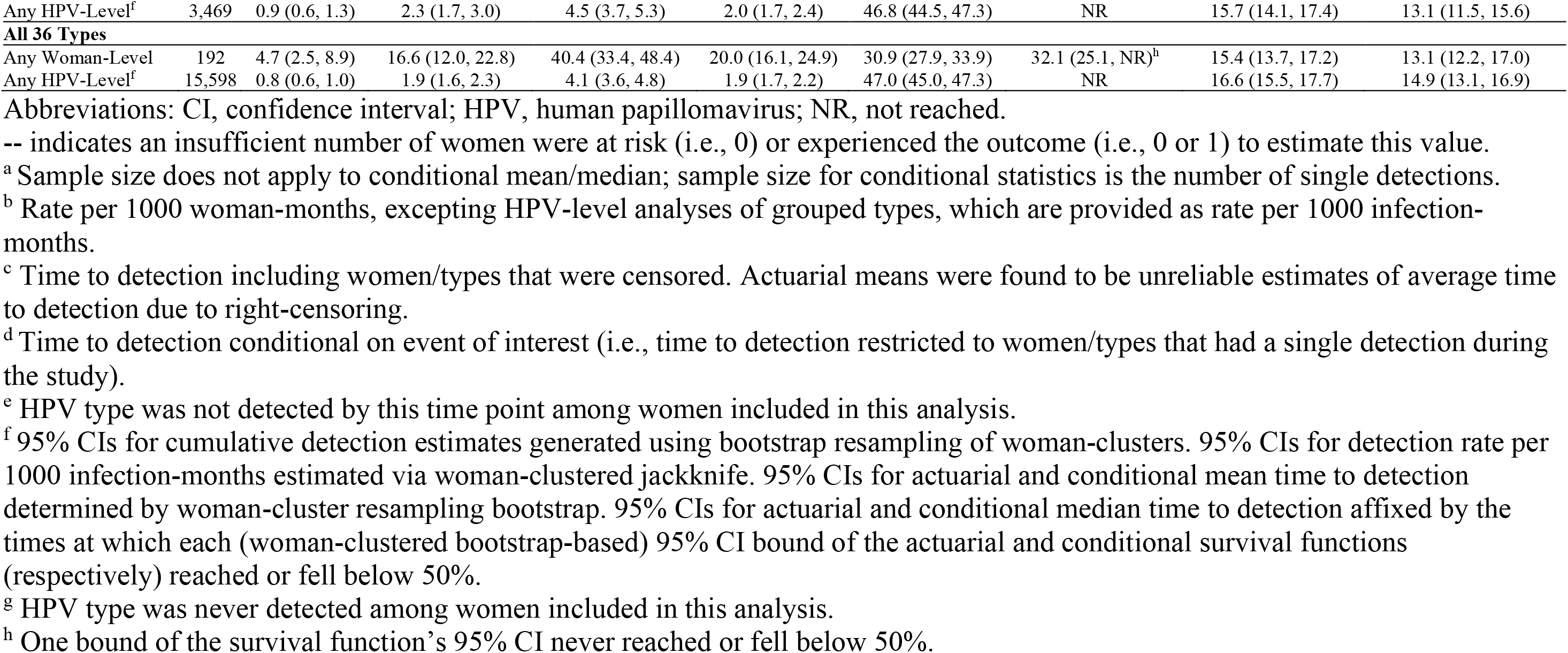
Single detection of incident infection for individual HPV types, grouped types at the woman-level, and grouped types at the HPV-level, by subgenus.

**Figure 2.**
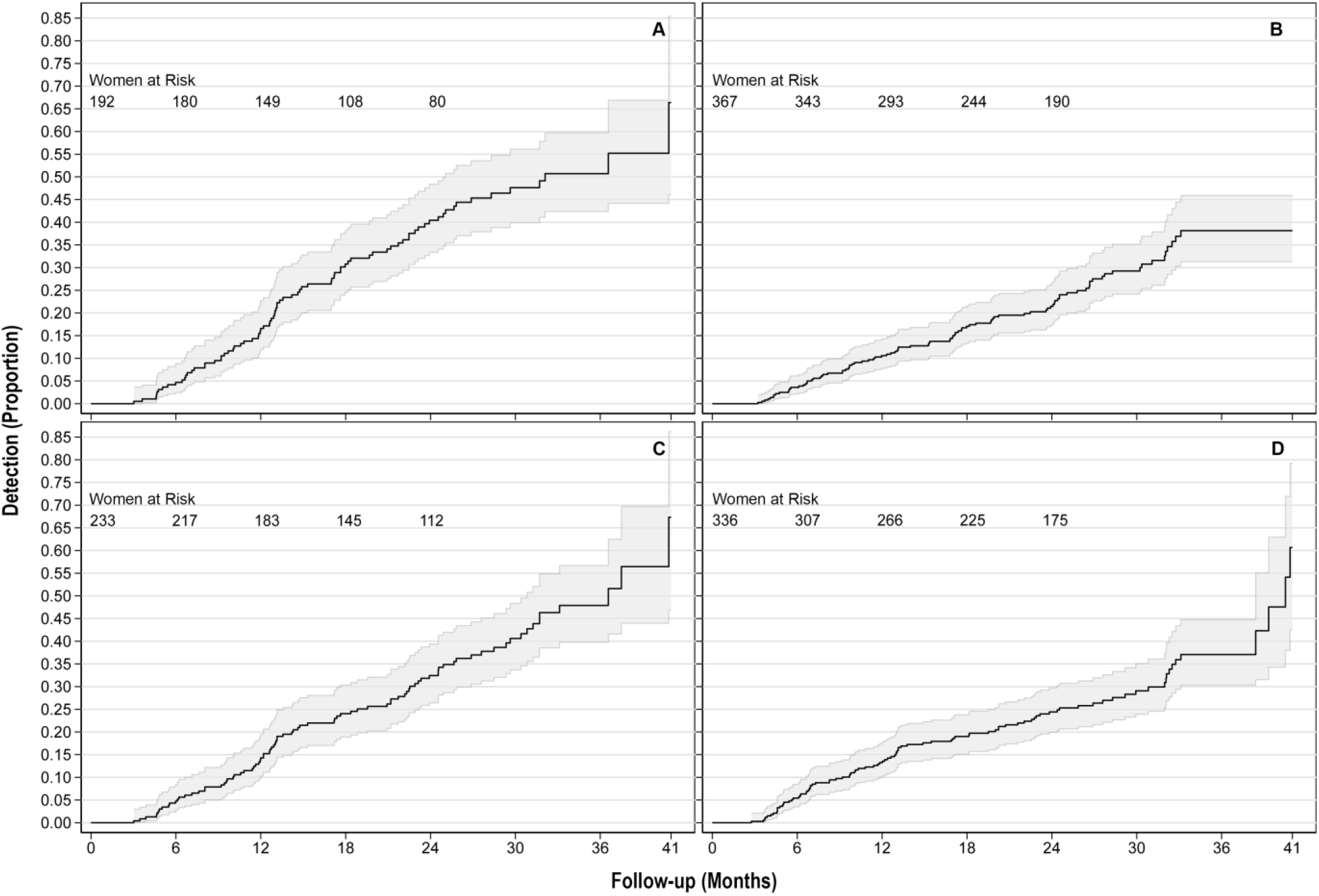
Single detection of incident infection with any (A) HPV type(s), (B) subgenus 1, (C) subgenus 2, and (D) subgenus 3 type(s), at the woman-level.

Tables 3 and 4 summarize liberal clearance analyses of present-at-baseline and incident infections, respectively. At the woman-level, the median times to clearance of all present-at-baseline and incident infections were 27.0, CI:25.0-32.7 and 22.3, CI:16.2-NR months, respectively (NR: CI bound Not Reached). At the HPV-level, the median time required to clear HPV infections of any type was similar between infections present at baseline (11.7, CI:10.3-12.6 months) and incident infections (12.6, CI:10.3-14.8 months). The rate of clearing infections of any HPV type per 1000 infection-months was different between infections present at baseline (61.3, CI:56.2-66.9) and incident infections (46.2, CI:41.1-52.0) at the HPV-level. This difference in rates was not observable at the woman-level (23.6 vs. 25.5 per 1000 woman-months). Figure 3 depicts, at the woman-level, survival functions for liberal clearance analyses; Figure S6 presents the same at the HPV-level. In HPV-level liberal clearance analyses, a larger proportion of incident infections (35.4%) were right censored compared to infections present at baseline (23.7%).

**Table 3.**
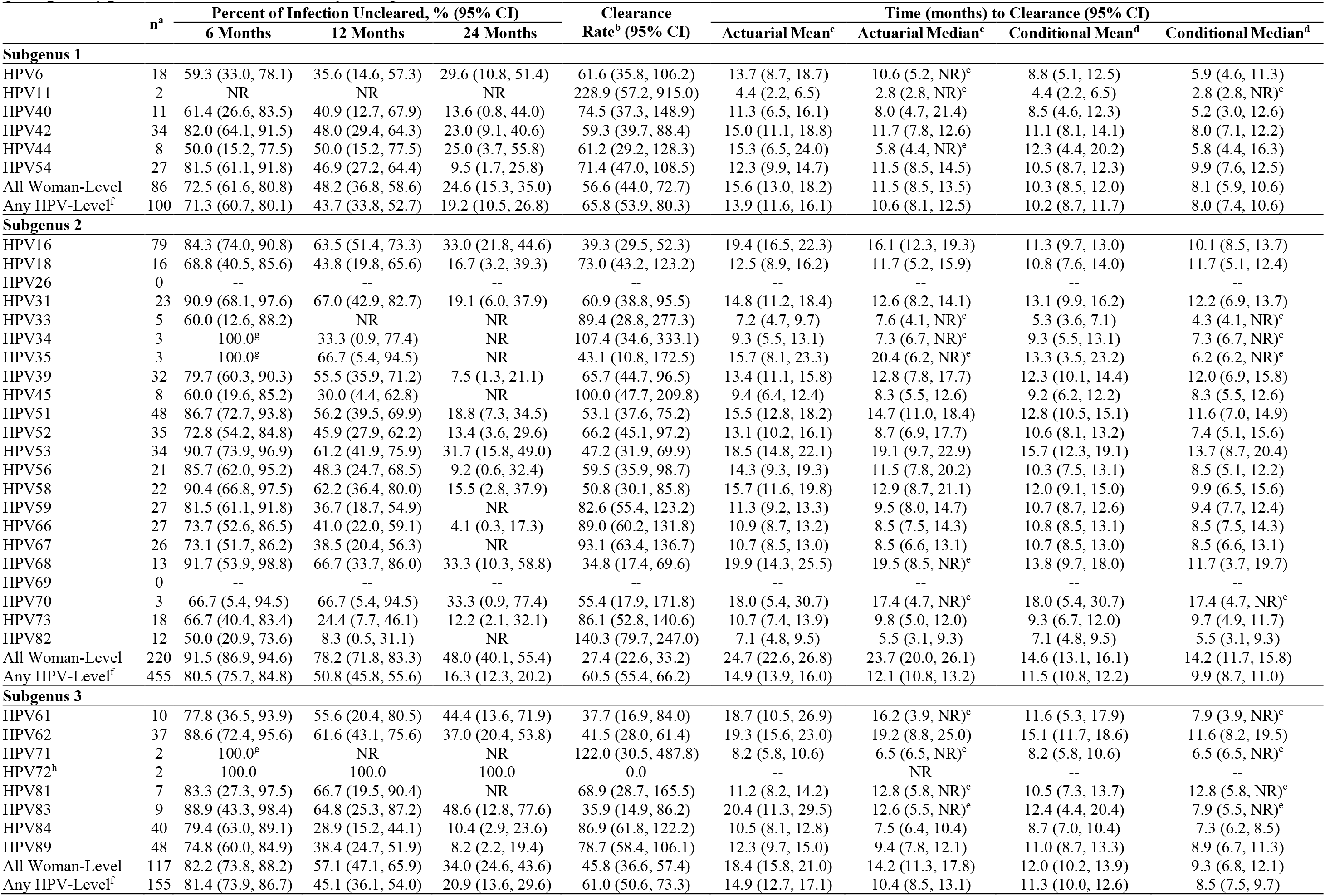
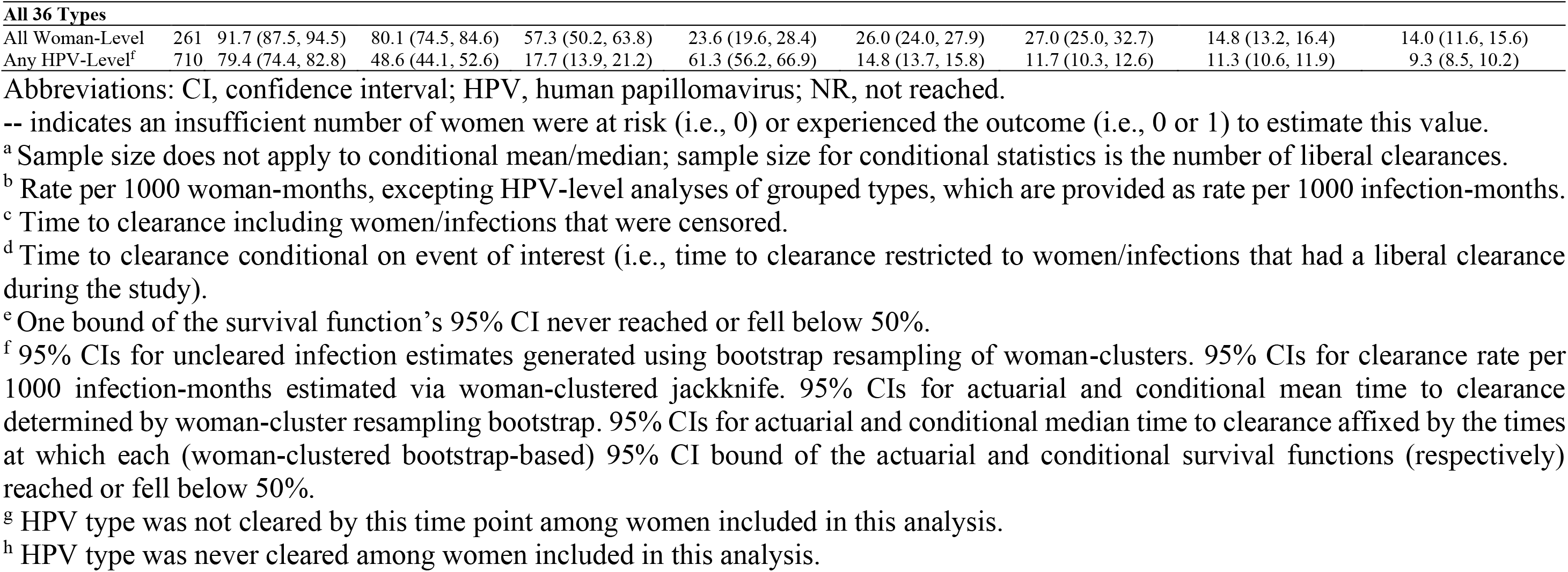
Liberal clearance of HPV infection present at baseline for individual HPV types, grouped types at the woman-level, and grouped types at the HPV-level, by subgenus.

|  | n<sup>a</sup> | Percent of Infection Uncleared, % (95% CI) |  |  | Clearance Rate<sup>b</sup> (95% CI) | Time (months) to Clearance (95% CI) |  |  |  |
| --- | --- | --- | --- | --- | --- | --- | --- | --- | --- |
|  |  | 6 Months | 12 Months | 24 Months |  | Actuarial Mean<sup>c</sup> | Actuarial Median<sup>c</sup> | Conditional Mean<sup>d</sup> | Conditional Median<sup>d</sup> |
| Subgenus 1 |  |  |  |  |  |  |  |  |  |
| HPV6 | 18 | 59.3 (33.0, 78.1) | 35.6 (14.6, 57.3) | 29.6 (10.8, 51.4) | 61.6 (35.8, 106.2) | 13.7 (8.7, 18.7) | 10.6 (5.2, NR)<sup>e</sup> | 8.8 (5.1, 12.5) | 5.9 (4.6, 11.3) |
| HPV11 | 2 | NR | NR | NR | 228.9 (57.2, 915.0) | 4.4 (2.2, 6.5) | 2.8 (2.8, NR)<sup>e</sup> | 4.4 (2.2, 6.5) | 2.8 (2.8, NR)<sup>e</sup> |
| HPV40 | 11 | 61.4 (26.6, 83.5) | 40.9 (12.7, 67.9) | 13.6 (0.8, 44.0) | 74.5 (37.3, 148.9) | 11.3 (6.5, 16.1) | 8.0 (4.7, 21.4) | 8.5 (4.6, 12.3) | 5.2 (3.0, 12.6) |
| HPV42 | 34 | 82.0 (64.1, 91.5) | 48.0 (29.4, 64.3) | 23.0 (9.1, 40.6) | 59.3 (39.7, 88.4) | 15.0 (11.1, 18.8) | 11.7 (7.8, 12.6) | 11.1 (8.1, 14.1) | 8.0 (7.1, 12.2) |
| HPV44 | 8 | 50.0 (15.2, 77.5) | 50.0 (15.2, 77.5) | 25.0 (3.7, 55.8) | 61.2 (29.2, 128.3) | 15.3 (6.5, 24.0) | 5.8 (4.4, NR)<sup>e</sup> | 12.3 (4.4, 20.2) | 5.8 (4.4, 16.3) |
| HPV54 | 27 | 81.5 (61.1, 91.8) | 46.9 (27.2, 64.4) | 9.5 (1.7, 25.8) | 71.4 (47.0, 108.5) | 12.3 (9.9, 14.7) | 11.5 (8.5, 14.5) | 10.5 (8.7, 12.3) | 9.9 (7.6, 12.5) |
| All Woman-Level | 86 | 72.5 (61.6, 80.8) | 48.2 (36.8, 58.6) | 24.6 (15.3, 35.0) | 56.6 (44.0, 72.7) | 15.6 (13.0, 18.2) | 11.5 (8.5, 13.5) | 10.3 (8.5, 12.0) | 8.1 (5.9, 10.6) |
| Any HPV-Level<sup>f</sup> | 100 | 71.3 (60.7, 80.1) | 43.7 (33.8, 52.7) | 19.2 (10.5, 26.8) | 65.8 (53.9, 80.3) | 13.9 (11.6, 16.1) | 10.6 (8.1, 12.5) | 10.2 (8.7, 11.7) | 8.0 (7.4, 10.6) |
| Subgenus 2 |  |  |  |  |  |  |  |  |  |
| HPV16 | 79 | 84.3 (74.0, 90.8) | 63.5 (51.4, 73.3) | 33.0 (21.8, 44.6) | 39.3 (29.5, 52.3) | 19.4 (16.5, 22.3) | 16.1 (12.3, 19.3) | 11.3 (9.7, 13.0) | 10.1 (8.5, 13.7) |
| HPV18 | 16 | 68.8 (40.5, 85.6) | 43.8 (19.8, 65.6) | 16.7 (3.2, 39.3) | 73.0 (43.2, 123.2) | 12.5 (8.9, 16.2) | 11.7 (5.2, 15.9) | 10.8 (7.6, 14.0) | 11.7 (5.1, 12.4) |
| HPV26 | 0 | -- | -- | -- | -- | -- | -- | -- | -- |
| HPV31 | 23 | 90.9 (68.1, 97.6) | 67.0 (42.9, 82.7) | 19.1 (6.0, 37.9) | 60.9 (38.8, 95.5) | 14.8 (11.2, 18.4) | 12.6 (8.2, 14.1) | 13.1 (9.9, 16.2) | 12.2 (6.9, 13.7) |
| HPV33 | 5 | 60.0 (12.6, 88.2) | NR | NR | 89.4 (28.8, 277.3) | 7.2 (4.7, 9.7) | 7.6 (4.1, NR)<sup>e</sup> | 5.3 (3.6, 7.1) | 4.3 (4.1, NR)<sup>e</sup> |
| HPV34 | 3 | 100.0<sup>g</sup> | 33.3 (0.9, 77.4) | NR | 107.4 (34.6, 333.1) | 9.3 (5.5, 13.1) | 7.3 (6.7, NR)<sup>e</sup> | 9.3 (5.5, 13.1) | 7.3 (6.7, NR)<sup>e</sup> |
| HPV35 | 3 | 100.0<sup>g</sup> | 66.7 (5.4, 94.5) | NR | 43.1 (10.8, 172.5) | 15.7 (8.1, 23.3) | 20.4 (6.2, NR)<sup>e</sup> | 13.3 (3.5, 23.2) | 6.2 (6.2, NR)<sup>e</sup> |
| HPV39 | 32 | 79.7 (60.3, 90.3) | 55.5 (35.9, 71.2) | 7.5 (1.3, 21.1) | 65.7 (44.7, 96.5) | 13.4 (11.1, 15.8) | 12.8 (7.8, 17.7) | 12.3 (10.1, 14.4) | 12.0 (6.9, 15.8) |
| HPV45 | 8 | 60.0 (19.6, 85.2) | 30.0 (4.4, 62.8) | NR | 100.0 (47.7, 209.8) | 9.4 (6.4, 12.4) | 8.3 (5.5, 12.6) | 9.2 (6.2, 12.2) | 8.3 (5.5, 12.6) |
| HPV51 | 48 | 86.7 (72.7, 93.8) | 56.2 (39.5, 69.9) | 18.8 (7.3, 34.5) | 53.1 (37.6, 75.2) | 15.5 (12.8, 18.2) | 14.7 (11.0, 18.4) | 12.8 (10.5, 15.1) | 11.6 (7.0, 14.9) |
| HPV52 | 35 | 72.8 (54.2, 84.8) | 45.9 (27.9, 62.2) | 13.4 (3.6, 29.6) | 66.2 (45.1, 97.2) | 13.1 (10.2, 16.1) | 8.7 (6.9, 17.7) | 10.6 (8.1, 13.2) | 7.4 (5.1, 15.6) |
| HPV53 | 34 | 90.7 (73.9, 96.9) | 61.2 (41.9, 75.9) | 31.7 (15.8, 49.0) | 47.2 (31.9, 69.9) | 18.5 (14.8, 22.1) | 19.1 (9.7, 22.9) | 15.7 (12.3, 19.1) | 13.7 (8.7, 20.4) |
| HPV56 | 21 | 85.7 (62.0, 95.2) | 48.3 (24.7, 68.5) | 9.2 (0.6, 32.4) | 59.5 (35.9, 98.7) | 14.3 (9.3, 19.3) | 11.5 (7.8, 20.2) | 10.3 (7.5, 13.1) | 8.5 (5.1, 12.2) |
| HPV58 | 22 | 90.4 (66.8, 97.5) | 62.2 (36.4, 80.0) | 15.5 (2.8, 37.9) | 50.8 (30.1, 85.8) | 15.7 (11.6, 19.8) | 12.9 (8.7, 21.1) | 12.0 (9.1, 15.0) | 9.9 (6.5, 15.6) |
| HPV59 | 27 | 81.5 (61.1, 91.8) | 36.7 (18.7, 54.9) | NR | 82.6 (55.4, 123.2) | 11.3 (9.2, 13.3) | 9.5 (8.0, 14.7) | 10.7 (8.7, 12.6) | 9.4 (7.7, 12.4) |
| HPV66 | 27 | 73.7 (52.6, 86.5) | 41.0 (22.0, 59.1) | 4.1 (0.3, 17.3) | 89.0 (60.2, 131.8) | 10.9 (8.7, 13.2) | 8.5 (7.5, 14.3) | 10.8 (8.5, 13.1) | 8.5 (7.5, 14.3) |
| HPV67 | 26 | 73.1 (51.7, 86.2) | 38.5 (20.4, 56.3) | NR | 93.1 (63.4, 136.7) | 10.7 (8.5, 13.0) | 8.5 (6.6, 13.1) | 10.7 (8.5, 13.0) | 8.5 (6.6, 13.1) |
| HPV68 | 13 | 91.7 (53.9, 98.8) | 66.7 (33.7, 86.0) | 33.3 (10.3, 58.8) | 34.8 (17.4, 69.6) | 19.9 (14.3, 25.5) | 19.5 (8.5, NR)<sup>e</sup> | 13.8 (9.7, 18.0) | 11.7 (3.7, 19.7) |
| HPV69 | 0 | -- | -- | -- | -- | -- | -- | -- | -- |
| HPV70 | 3 | 66.7 (5.4, 94.5) | 66.7 (5.4, 94.5) | 33.3 (0.9, 77.4) | 55.4 (17.9, 171.8) | 18.0 (5.4, 30.7) | 17.4 (4.7, NR)<sup>e</sup> | 18.0 (5.4, 30.7) | 17.4 (4.7, NR)<sup>e</sup> |
| HPV73 | 18 | 66.7 (40.4, 83.4) | 24.4 (7.7, 46.1) | 12.2 (2.1, 32.1) | 86.1 (52.8, 140.6) | 10.7 (7.4, 13.9) | 9.8 (5.0, 12.0) | 9.3 (6.7, 12.0) | 9.7 (4.9, 11.7) |
| HPV82 | 12 | 50.0 (20.9, 73.6) | 8.3 (0.5, 31.1) | NR | 140.3 (79.7, 247.0) | 7.1 (4.8, 9.5) | 5.5 (3.1, 9.3) | 7.1 (4.8, 9.5) | 5.5 (3.1, 9.3) |
| All Woman-Level | 220 | 91.5 (86.9, 94.6) | 78.2 (71.8, 83.3) | 48.0 (40.1, 55.4) | 27.4 (22.6, 33.2) | 24.7 (22.6, 26.8) | 23.7 (20.0, 26.1) | 14.6 (13.1, 16.1) | 14.2 (11.7, 15.8) |
| Any HPV-Level<sup>f</sup> | 455 | 80.5 (75.7, 84.8) | 50.8 (45.8, 55.6) | 16.3 (12.3, 20.2) | 60.5 (55.4, 66.2) | 14.9 (13.9, 16.0) | 12.1 (10.8, 13.2) | 11.5 (10.8, 12.2) | 9.9 (8.7, 11.0) |
| Subgenus 3 |  |  |  |  |  |  |  |  |  |
| HPV61 | 10 | 77.8 (36.5, 93.9) | 55.6 (20.4, 80.5) | 44.4 (13.6, 71.9) | 37.7 (16.9, 84.0) | 18.7 (10.5, 26.9) | 16.2 (3.9, NR)<sup>e</sup> | 11.6 (5.3, 17.9) | 7.9 (3.9, NR)<sup>e</sup> |
| HPV62 | 37 | 88.6 (72.4, 95.6) | 61.6 (43.1, 75.6) | 37.0 (20.4, 53.8) | 41.5 (28.0, 61.4) | 19.3 (15.6, 23.0) | 19.2 (8.8, 25.0) | 15.1 (11.7, 18.6) | 11.6 (8.2, 19.5) |
| HPV71 | 2 | 100.0<sup>g</sup> | NR | NR | 122.0 (30.5, 487.8) | 8.2 (5.8, 10.6) | 6.5 (6.5, NR)<sup>e</sup> | 8.2 (5.8, 10.6) | 6.5 (6.5, NR)<sup>e</sup> |
| HPV72<sup>h</sup> | 2 | 100.0 | 100.0 | 100.0 | 0.0 | -- | NR | -- | -- |
| HPV81 | 7 | 83.3 (27.3, 97.5) | 66.7 (19.5, 90.4) | NR | 68.9 (28.7, 165.5) | 11.2 (8.2, 14.2) | 12.8 (5.8, NR)<sup>e</sup> | 10.5 (7.3, 13.7) | 12.8 (5.8, NR)<sup>e</sup> |
| HPV83 | 9 | 88.9 (43.3, 98.4) | 64.8 (25.3, 87.2) | 48.6 (12.8, 77.6) | 35.9 (14.9, 86.2) | 20.4 (11.3, 29.5) | 12.6 (5.5, NR)<sup>e</sup> | 12.4 (4.4, 20.4) | 7.9 (5.5, NR)<sup>e</sup> |
| HPV84 | 40 | 79.4 (63.0, 89.1) | 28.9 (15.2, 44.1) | 10.4 (2.9, 23.6) | 86.9 (61.8, 122.2) | 10.5 (8.1, 12.8) | 7.5 (6.4, 10.4) | 8.7 (7.0, 10.4) | 7.3 (6.2, 8.5) |
| HPV89 | 48 | 74.8 (60.0, 84.9) | 38.4 (24.7, 51.9) | 8.2 (2.2, 19.4) | 78.7 (58.4, 106.1) | 12.3 (9.7, 15.0) | 9.4 (7.8, 12.1) | 11.0 (8.7, 13.3) | 8.9 (6.7, 11.3) |
| All Woman-Level | 117 | 82.2 (73.8, 88.2) | 57.1 (47.1, 65.9) | 34.0 (24.6, 43.6) | 45.8 (36.6, 57.4) | 18.4 (15.8, 21.0) | 14.2 (11.3, 17.8) | 12.0 (10.2, 13.9) | 9.3 (6.8, 12.1) |
| Any HPV-Level<sup>f</sup> | 155 | 81.4 (73.9, 86.7) | 45.1 (36.1, 54.0) | 20.9 (13.6, 29.6) | 61.0 (50.6, 73.3) | 14.9 (12.7, 17.1) | 10.4 (8.5, 13.1) | 11.3 (10.0, 12.6) | 8.5 (7.5, 9.7) |
| **All 36 Types** |  |  |  |  |  |  |  |  |  |
| All Woman-Level | 261 | 91.7 (87.5, 94.5) | 80.1 (74.5, 84.6) | 57.3 (50.2, 63.8) | 23.6 (19.6, 28.4) | 26.0 (24.0, 27.9) | 27.0 (25.0, 32.7) | 14.8 (13.2, 16.4) | 14.0 (11.6, 15.6) |
| Any HPV-Level<sup>f</sup> | 710 | 79.4 (74.4, 82.8) | 48.6 (44.1, 52.6) | 17.7 (13.9, 21.2) | 61.3 (56.2, 66.9) | 14.8 (13.7, 15.8) | 11.7 (10.3, 12.6) | 11.3 (10.6, 11.9) | 9.3 (8.5, 10.2) |

Abbreviations: CI, confidence interval; HPV, human papillomavirus; NR, not reached.
-- indicates an insufficient number of women were at risk (i.e., 0) or experienced the outcome (i.e., 0 or 1) to estimate this value.
<sup>a</sup> Sample size does not apply to conditional mean/median; sample size for conditional statistics is the number of liberal clearances.
<sup>b</sup> Rate per 1000 woman-months, excepting HPV-level analyses of grouped types, which are provided as rate per 1000 infection-months.
<sup>c</sup> Time to clearance including women/infections that were censored.
<sup>d</sup> Time to clearance conditional on event of interest (i.e., time to clearance restricted to women/infections that had a liberal clearance during the study).
<sup>e</sup> One bound of the survival function's 95% CI never reached or fell below 50%.
<sup>f</sup> 95% CIs for uncleared infection estimates generated using bootstrap resampling of woman-clusters. 95% CIs for clearance rate per 1000 infection-months estimated via woman-clustered jackknife. 95% CIs for actuarial and conditional mean time to clearance determined by woman-cluster resampling bootstrap. 95% CIs for actuarial and conditional median time to clearance affixed by the times at which each (woman-clustered bootstrap-based) 95% CI bound of the actuarial and conditional survival functions (respectively) reached or fell below 50%.
<sup>g</sup> HPV type was not cleared by this time point among women included in this analysis.
<sup>h</sup> HPV type was never cleared among women included in this analysis.

**Table 4.** Liberal clearance of incident infection for individual HPV types, grouped types at the woman-level, and grouped types at the HPV-level, by subgenus.

|  | n<sup>a</sup> | Percent of Infection Uncleared, % (95% CI) |  |  | Clearance Rate<sup>b</sup> (95% CI) | Time (months) to Clearance (95% CI) |  |  |  |
| --- | --- | --- | --- | --- | --- | --- | --- | --- | --- |
|  |  | 6 Months | 12 Months | 24 Months |  | Actuarial Mean<sup>c</sup> | Actuarial Median<sup>c</sup> | Conditional Mean<sup>d</sup> | Conditional Median<sup>d</sup> |
| Subgenus 1 |  |  |  |  |  |  |  |  |  |
| HPV6 | 32 | 79.4 (59.8, 90.2) | 35.2 (16.8, 54.2) | NR | 66.4 (41.8, 105.3) | 12.1 (9.3, 14.8) | 9.3 (7.6, 12.8) | 8.4 (6.5, 10.2) | 7.6 (5.3, 9.3) |
| HPV11 | 1 | NR | NR | NR | 172.9 (24.4, 1,227.7) | -- | NR | -- | NR |
| HPV40 | 14 | 85.7 (53.9, 96.2) | 47.6 (20.3, 70.8) | NR | 73.9 (40.9, 133.4) | 11.5 (9.1, 13.8) | 10.2 (6.1, 16.2) | 10.5 (7.9, 13.0) | 9.9 (5.2, 15.1) |
| HPV42 | 34 | 82.0 (64.3, 91.5) | 52.1 (33.3, 67.9) | 23.1 (9.1, 40.9) | 55.1 (36.3, 83.7) | 14.7 (11.4, 18.0) | 12.4 (8.6, 16.0) | 9.7 (8.0, 11.4) | 8.6 (5.7, 12.4) |
| HPV44 | 4 | 100.0<sup>e</sup> | 33.3 (0.9, 77.4) | NR | 74.2 (23.9, 230.2) | 11.3 (5.7, 16.9) | 8.5 (7.2, NR)<sup>f</sup> | 11.3 (5.7, 16.9) | 8.5 (7.2, NR)<sup>f</sup> |
| HPV54 | 19 | 89.5 (64.1, 97.3) | 44.2 (17.5, 68.2) | 22.1 (1.6, 57.5) | 50.6 (27.2, 94.0) | 15.0 (10.8, 19.3) | 10.6 (7.9, NR)<sup>f</sup> | 10.8 (6.5, 15.1) | 8.0 (3.0, 10.6) |
| All Woman-Level | 69 | 86.8 (76.1, 92.9) | 55.0 (40.9, 67.1) | 19.9 (6.5, 38.6) | 45.6 (32.7, 63.5) | 15.6 (13.0, 18.2) | 14.3 (9.9, 17.6) | 9.8 (8.2, 11.4) | 8.7 (7.3, 10.0) |
| Any HPV-Level<sup>g</sup> | 104 | 83.5 (74.8, 89.4) | 52.3 (41.1, 62.1) | 34.7 (24.2, 44.8) | 43.4 (33.6, 56.4) | 18.5 (15.4, 21.1) | 13.1 (9.9, 17.6) | 9.6 (8.6, 10.9) | 8.5 (7.6, 9.9) |
| Subgenus 2 |  |  |  |  |  |  |  |  |  |
| HPV16 | 22 | 83.3 (56.8, 94.3) | 67.3 (36.6, 85.6) | NR | 36.5 (17.4, 76.6) | 14.0 (10.7, 17.3) | 15.3 (6.7, NR)<sup>f</sup> | 8.5 (5.7, 11.4) | 6.7 (4.8, 13.0) |
| HPV18 | 10 | 80.0 (40.9, 94.6) | 50.0 (18.4, 75.3) | 50.0 (18.4, 75.3) | 34.7 (14.5, 83.4) | 18.6 (11.4, 25.8) | 10.2 (4.6, NR)<sup>f</sup> | 7.1 (5.4, 8.8) | 7.4 (4.6, NR)<sup>f</sup> |
| HPV26 | 0 | -- | -- | -- | -- | -- | -- | -- | -- |
| HPV31 | 19 | 78.0 (51.5, 91.1) | 36.0 (13.6, 59.3) | NR | 69.1 (39.2, 121.7) | 11.3 (8.4, 14.1) | 9.8 (6.8, NR)<sup>f</sup> | 8.6 (6.2, 10.9) | 8.0 (4.6, 10.0) |
| HPV33 | 4 | 50.0 (5.8, 84.5) | NR | NR | 153.1 (57.5, 408.0) | 6.5 (4.5, 8.5) | 5.5 (3.7, NR)<sup>f</sup> | 6.5 (4.5, 8.5) | 5.5 (3.7, NR)<sup>f</sup> |
| HPV34 | 2 | 50.0 (0.6, 91.0) | NR | NR | 168.6 (42.2, 674.2) | 5.9 (5.8, 6.0) | 5.8 (5.8, NR)<sup>f</sup> | 5.9 (5.8, 6.0) | 5.8 (5.8, NR)<sup>f</sup> |
| HPV35 | 4 | 75.0 (12.8, 96.1) | 75.0 (12.8, 96.1) | NR | 55.3 (17.8, 171.5) | 16.6 (9.7, 23.5) | 18.5 (5.0, NR)<sup>f</sup> | 15.4 (6.9, 23.8) | 18.5 (5.0, NR)<sup>f</sup> |
| HPV39 | 19 | 89.5 (64.1, 97.3) | 57.0 (29.7, 77.1) | 19.0 (3.2, 44.9) | 54.0 (29.9, 97.5) | 14.1 (9.5, 18.7) | 12.6 (8.1, 14.8) | 9.6 (7.6, 11.6) | 8.5 (5.6, 12.6) |
| HPV45 | 8 | 75.0 (31.5, 93.1) | 20.8 (1.0, 58.6) | NR | 90.7 (40.8, 201.9) | 9.4 (5.3, 13.4) | 7.6 (4.7, NR)<sup>f</sup> | 8.6 (4.9, 12.3) | 6.2 (4.7, NR)<sup>f</sup> |
| HPV51 | 23 | 86.7 (64.3, 95.5) | 47.2 (23.4, 67.8) | 12.6 (1.0, 39.3) | 61.8 (37.3, 102.6) | 13.7 (9.9, 17.5) | 11.3 (6.3, 16.3) | 11.1 (7.6, 14.7) | 9.4 (4.9, 15.1) |
| HPV52 | 19 | 89.2 (63.2, 97.2) | 50.2 (26.1, 70.2) | 13.7 (1.1, 41.7) | 59.1 (34.3, 101.8) | 13.6 (9.9, 17.3) | 12.6 (6.8, 23.0) | 10.0 (7.1, 12.8) | 9.0 (6.2, 12.6) |
| HPV53 | 25 | 78.7 (56.1, 90.6) | 64.1 (40.5, 80.3) | NR | 56.0 (33.2, 94.5) | 13.7 (10.5, 16.9) | 14.1 (6.4, 18.4) | 10.0 (6.8, 13.3) | 6.4 (3.7, 14.9) |
| HPV56 | 16 | 85.7 (53.9, 96.2) | 39.6 (12.8, 65.9) | NR | 70.4 (37.9, 130.8) | 10.7 (8.2, 13.2) | 8.6 (6.3, 15.5) | 9.7 (7.2, 12.1) | 8.2 (5.3, 12.7) |
| HPV58 | 13 | 92.3 (56.6, 98.9) | 92.3 (56.6, 98.9) | 20.2 (0.9, 58.0) | 31.8 (14.3, 70.7) | 21.5 (15.4, 27.6) | 22.3 (12.2, NR)<sup>f</sup> | 19.1 (11.6, 26.5) | 18.2 (4.5, NR)<sup>f</sup> |
| HPV59 | 18 | 70.8 (43.5, 86.7) | 27.0 (6.9, 52.7) | 27.0 (6.9, 52.7) | 67.5 (37.4, 121.9) | 13.7 (7.8, 19.5) | 9.4 (5.1, NR)<sup>f</sup> | 9.1 (4.8, 13.3) | 7.1 (4.6, 9.9) |
| HPV66 | 35 | 73.2 (54.8, 85.1) | 33.1 (16.6, 50.6) | NR | 91.8 (63.0, 133.9) | 9.7 (7.9, 11.4) | 8.8 (6.2, 13.1) | 8.7 (7.0, 10.4) | 7.6 (5.3, 10.2) |
| HPV67 | 25 | 88.0 (67.3, 96.0) | 37.8 (19.2, 56.3) | NR | 80.4 (52.4, 123.3) | 10.9 (9.0, 12.8) | 8.5 (7.2, 14.0) | 9.8 (8.0, 11.7) | 8.3 (6.9, 11.3) |
| HPV68 | 6 | 80.0 (20.4, 96.9) | 26.7 (1.0, 68.6) | NR | 57.7 (18.6, 179.0) | 11.0 (8.0, 14.0) | 11.5 (5.6, NR)<sup>f</sup> | 9.2 (6.2, 12.1) | 10.5 (5.6, NR)<sup>f</sup> |
| HPV69 | 0 | -- | -- | -- | -- | -- | -- | -- | -- |
| HPV70 | 5 | 80.0 (20.4, 96.9) | 40.0 (5.2, 75.3) | 40.0 (5.2, 75.3) | 65.6 (24.6, 174.7) | 14.4 (5.8, 23.0) | 7.3 (5.6, NR)<sup>f</sup> | 11.4 (2.8, 19.9) | 6.2 (5.6, NR)<sup>f</sup> |
| HPV73 | 21 | 81.0 (56.9, 92.4) | 40.7 (18.7, 61.8) | NR | 79.7 (48.8, 130.1) | 11.0 (8.9, 13.2) | 9.6 (7.5, 16.1) | 10.0 (7.8, 12.3) | 8.8 (5.3, 15.8) |
| HPV82 | 9 | 77.8 (36.5, 93.9) | 22.2 (3.4, 51.3) | NR | 81.2 (38.7, 170.3) | 9.6 (7.1, 12.1) | 9.3 (3.8, NR)<sup>f</sup> | 7.8 (6.3, 9.4) | 8.9 (3.8, 9.4) |
| All Woman-Level | 68 | 87.7 (76.8, 93.6) | 57.1 (42.8, 69.2) | 18.4 (6.3, 35.5) | 47.2 (34.1, 65.5) | 17.2 (13.4, 21.0) | 13.5 (9.4, 18.5) | 10.1 (8.4, 11.9) | 8.3 (6.4, 11.3) |
| Any HPV-Level<sup>g</sup> | 303 | 81.7 (76.1, 86.2) | 51.9 (45.0, 57.7) | 31.0 (24.1, 36.5) | 47.1 (39.9, 55.5) | 19.1 (15.9, 21.2) | 12.7 (9.4, 15.1) | 9.7 (8.9, 10.6) | 8.1 (7.3, 8.9) |
| Subgenus 3 |  |  |  |  |  |  |  |  |  |
| HPV61 | 16 | 81.3 (52.5, 93.5) | 48.8 (17.1, 74.7) | NR | 42.8 (20.4, 89.9) | 14.5 (10.0, 18.9) | 11.9 (7.9, NR)<sup>f</sup> | 8.0 (5.3, 10.8) | 7.9 (3.0, 11.9) |
| HPV62 | 25 | 79.5 (57.5, 90.9) | 46.0 (24.3, 65.3) | 12.6 (1.1, 38.9) | 65.7 (40.8, 105.7) | 13.1 (9.4, 16.7) | 11.7 (6.8, 14.4) | 10.4 (7.4, 13.3) | 8.2 (5.7, 12.2) |
| HPV71 | 0 | -- | -- | -- | -- | -- | -- | -- | -- |
| HPV72 | 0 | -- | -- | -- | -- | -- | -- | -- | -- |
| HPV81 | 2 | 100.0<sup>e</sup> | 100.0<sup>e</sup> | NR | 44.8 (6.3, 318.2) | -- | 12.2 (NR, NR)<sup>f</sup> | -- | NR |
| HPV83 | 7 | 100.0<sup>e</sup> | 17.1 (0.8, 52.6) | NR | 82.5 (34.4, 198.3) | 9.1 (7.4, 10.8) | 8.8 (6.2, NR)<sup>f</sup> | 8.2 (7.0, 9.5) | 8.5 (6.2, NR)<sup>f</sup> |
| HPV84 | 34 | 63.9 (45.2, 77.7) | 37.0 (19.7, 54.4) | 14.8 (4.1, 31.9) | 79.2 (53.5, 117.1) | 11.5 (8.6, 14.5) | 8.5 (5.6, 15.1) | 8.8 (6.4, 11.1) | 6.1 (4.6, 9.9) |
| HPV89 | 43 | 88.0 (73.6, 94.8) | 51.4 (34.7, 65.8) | 20.9 (6.7, 40.5) | 57.5 (39.7, 83.3) | 14.4 (11.6, 17.2) | 12.9 (8.1, 20.1) | 11.3 (8.7, 13.9) | 8.1 (6.7, 11.3) |
| All Woman-Level | 71 | 80.1 (68.7, 87.7) | 59.1 (45.6, 70.3) | 26.7 (13.2, 42.3) | 51.0 (37.4, 69.6) | 15.3 (12.9, 17.7) | 14.4 (10.8, 19.7) | 10.4 (8.2, 12.5) | 6.8 (5.7, 11.8) |
| Any HPV-Level<sup>g</sup> | 127 | 80.3 (72.4, 85.6) | 50.9 (40.3, 58.8) | 35.0 (24.5, 42.0) | 46.6 (37.7, 57.7) | 17.3 (14.8, 19.4) | 12.2 (9.7, 15.1) | 9.9 (8.6, 11.4) | 8.0 (6.7, 9.6) |
| **All 36 Types** |  |  |  |  |  |  |  |  |  |
| All Woman-Level | 65 | 93.5 (83.6, 97.5) | 74.1 (60.1, 83.9) | 39.8 (19.2, 59.7) | 25.5 (16.8, 38.7) | 24.2 (19.6, 28.9) | 22.3 (16.2, NR)<sup>f</sup> | 11.1 (8.7, 13.5) | 8.7 (6.4, 13.5) |
| Any HPV-Level<sup>g</sup> | 534 | 81.7 (77.1, 86.0) | 51.7 (46.6, 56.4) | 32.7 (27.4, 36.8) | 46.2 (41.1, 52.0) | 19.3 (16.9, 20.8) | 12.6 (10.3, 14.8) | 9.8 (9.1, 10.5) | 8.2 (7.6, 8.8) |

Abbreviations: CI, confidence interval; HPV, human papillomavirus; NR, not reached.
-- indicates an insufficient number of women were at risk (i.e., 0) or experienced the outcome (i.e., 0 or 1) to estimate this value.
<sup>a</sup> Sample size does not apply to conditional mean/median; sample size for conditional statistics is the number of liberal clearances.
<sup>b</sup> Rate per 1000 woman-months, excepting HPV-level analyses of grouped types, which are provided as rate per 1000 infection-months.
<sup>c</sup> Time to clearance including women/infections that were censored.
<sup>d</sup> Time to clearance conditional on event of interest (i.e., time to clearance restricted to women/infections that had a liberal clearance during the study).
<sup>e</sup> HPV type was not cleared by this time point among women included in this analysis.
<sup>f</sup> Bound(s) of the survival function's 95% CI never reached or fell below 50%.
<sup>g</sup> 95% CIs for uncleared infection estimates generated using bootstrap resampling of woman-clusters. 95% CIs for clearance rate per 1000 infection-months estimated via woman-clustered jackknife. 95% CIs for actuarial and conditional mean time to clearance determined by woman-cluster resampling bootstrap. 95% CIs for actuarial and conditional median time to clearance affixed by the times at which each (woman-clustered bootstrap-based) 95% CI bound of the actuarial and conditional survival functions (respectively) reached or fell below 50%.

**Figure 3.**
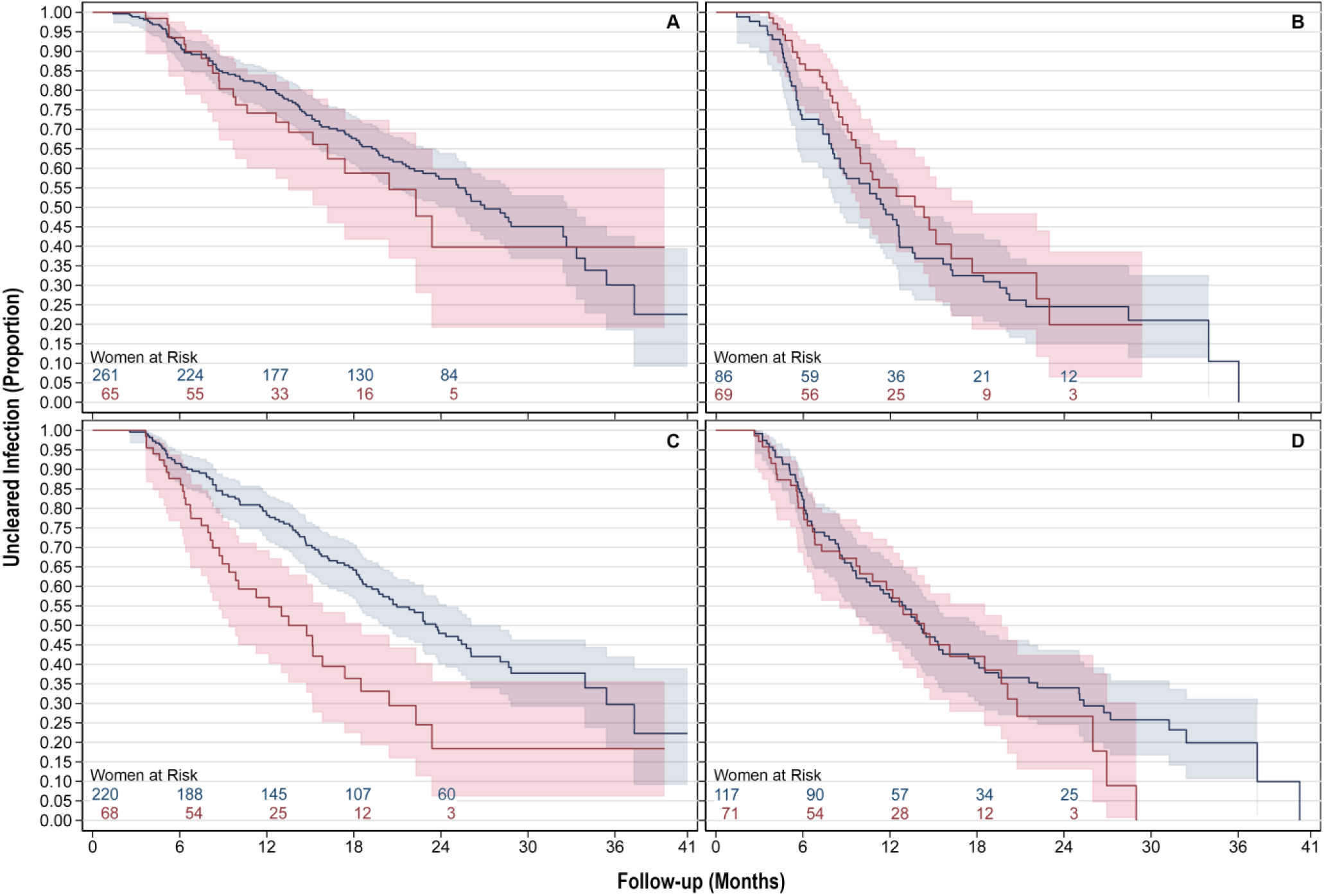
Liberal clearance of all (A) HPV type(s), (B) subgenus 1, (C) subgenus 2, and (D) subgenus 3 type(s) for infections present at baseline (blue) and incident infections (red), at the woman-level.

At the woman-level, among infections present at baseline, we observed a lower clearance rate per 1000 woman-months (27.4, CI:22.6-33.2), and a longer median time to clearance (23.7, CI:20.0-26.1 months) of all subgenus 2 infections compared to subgenera 1 (rate 56.6, CI:44.0-72.7; median 11.5, CI:8.5-13.5 months) and 3 (rate 45.8, CI:36.6-57.4; median 14.2, CI:11.3-17.8 months). Among incident infections, the clearance rate per 1000 woman-months (47.2, CI:34.1-65.5), and median time to clearance (13.5, CI:9.4-18.5 months) of all subgenus 2 infections were similar to those of subgenera 1 (rate 45.6, CI:32.7-63.5; median 14.3, CI:9.9-17.6 months) and 3 (rate 51.0, CI:37.4-69.6; median 14.4, CI:10.8-19.7 months).

At the HPV-level, however, there was homogeny within infections present at baseline and within incident infections when estimating the time to clearance of subgenus 2 infections compared to subgenera 1 and 3 infections. Amongst infections present at baseline, rates of clearing any infection of subgenera 1, 2, and 3 were 65.8, CI:53.9-80.3; 60.5, CI:55.4-66.2; and 61.0, CI:50.6-73.3 per 1000 infection-months, respectively. The corresponding median infection durations were 10.6, CI:8.1-12.5; 12.1, CI:10.8-13.2; and 10.4, CI:8.5-13.1 months, respectively. Amongst incident infections, rates of clearing any infection of subgenera 1, 2, and 3 were 43.4, CI:33.6-56.4; 47.1, CI:39.9-55.5; and 46.6, CI:37.7-57.7 per 1000 infection-months, respectively. The corresponding median infection durations were 13.1, CI:9.9-17.6; 12.7, CI:9.4-15.1; and 12.2, CI:9.7-15.1 months, respectively. The percent of right-censored infections in HPV-level grouped analyses was similar for subgenera 1, 2, and 3 within analyses of infections present-at-baseline (24.0%, 23.7%, and 23.2%, respectively) and within analyses of incident infections (37.5%, 35.0%, and 34.6%, respectively). Tables S2 and S3, alongside Figures S7 and S8 reproduce the aforementioned results for conservative clearance analyses.

Exponentially-extended means (calculated using a survival function extended to 0) are presented in Tables S4 (time to detection) and S5 (time to clearance). The average differences between exponentially-extended and actuarial mean times to single detection were 1538.7 months for individual HPV types, 28.4 months for grouped types at the woman-level, and 363.5 months for grouped types at the HPV-level, respectively. These large average differences suggest the mean time to detection was unreliable. For liberal clearance analyses, the corresponding average differences were 1.4, 3.1, and 0.2 months for infections present at baseline and 1.7, 6.2, and 7.3 months for incident infections. These small average differences suggest the mean time to clearance was reliable.

## DISCUSSION

This study described the natural history of vaginal HPV infections in young women who recently initiated a new sexual relationship. We found a high prevalence, high detection rate, low clearance rate, and long average duration of HPV16 infections compared to other HPV types, consistent with previous studies [11, 12, 14-16, 18-21]. We observed a higher woman-level rate of detection for any HPV type compared to pooled rates in women under 30 years of age (20.0 vs. 15.6 detections per 1000 woman-months, respectively) [12]. By 24 months, we estimated a similar cumulative detection of any HPV to those reached by 24 to 36 months in studies of similarly-aged or slightly younger women [15, 20, 21] and by 12 months in a study of slightly older young women [14]. 24-month cumulative detection and rate of detection in this cohort are comparable to a previous study of college-age women in Montréal [19]. The most likely explanation for the observed high rates of HPV detection in this cohort is that all women had recently begun a new sexual relationship at enrollment. A new sexual partner represents a potential for exposure to new HPV infection(s). In support of this explanation, a similar cumulative detection rate has been observed by 24 months in women with only one partner post-sexual debut [13]. Testing for many HPV types may also have played a role in the observed high detection rate.

The woman-level rate of incident subgenus 2 (i.e., oncogenic) infection detection was higher than the corresponding rates for subgenera 1 and 3. This finding is consistent with several studies observing elevated detection rates among high-risk types [12]. While we did not use the traditional high-risk vs. low-risk HPV grouping scheme, comparing subgenus 2 types to subgenera 1 and 3 types (i.e., high oncogenic risk types vs. low oncogenic risk & commensal types) is a viable analogue based on biological rationale.

Our woman-level estimates of median time to clearance of all infections present at baseline (27.0 months) and clearance of all incident infections (22.3 months) were larger than a meta-analyzed estimate based on studies that included prevalent and/or incident infections (9.8 months) [11]. In particular, it took women in our study longer, on average, to clear all incident infections compared to an earlier cohort of college-age women in Montréal [19]. Follow-up intervals in the present cohort were spaced similarly to those of the earlier study per protocol (4-6 months vs. 6 months, respectively). However, three-quarters or more women attended follow-up visits late in the present cohort, so interval censoring may have artificially prolonged the time to infection clearance.

In HPV-level analyses, the median duration of infection with any HPV type was similar between incident and present-at-baseline infections, though the clearance rate was higher for those present at baseline. However, our analysis of incident infections was more prone to right-censoring than our analysis of infections present at baseline, so we cannot rule out the possibility that women were under observation for an inadequate period to clear incident infections, artificially lowering the corresponding clearance rate estimate.

Among women with infections at baseline, we observed a lower woman-level clearance rate and longer median time to clearance of all subgenus 2 infections compared to subgenera 1 and 3. A meta-analysed estimate [11], and several longitudinal studies specific to young women [14, 17-19] corroborate lower woman-level clearance rates for high-risk vs. low-risk HPV types.

A lower rate of, and longer median time to clearance might suggest that subgenus 2 (i.e., oncogenic) infections are more persistent than infections belonging to other subgenera. However, woman-level analyses are limited in their ability to accurately estimate infection persistence. Woman-level clearance events are only counted when a woman tests negative for all HPV types within the grouping of interest. Assuming the null hypothesis, *Subgenus 2 infections are no more persistent than infections of other subgenera*, clearing (up to) twenty-two subgenus 2 infections is of lower probability than clearing (up to) six subgenus 1, or eight subgenus 3 infections. Past studies have generally genotyped more high- than low-risk HPVs, which creates a similar conundrum. In contrast to woman-level analyses, HPV-level analyses treat the clearance of each unique infection as a separate event. This is a more biologically informed paradigm for understanding the natural history of individual HPV infections.

In our HPV-level analyses, we did not observe systematic differences in the persistence of high oncogenic risk subgenus 2 infections compared to low oncogenic risk subgenera 1 and 3 infections, as indicated by clearance rate and median time to clearance. This was the case for both incident and present-at-baseline infections. It is unlikely that these results are an artefact of disproportional censoring, since subgenus 2 infections were no more right censor-prone than infections belonging to other subgenera. Altogether, our woman-level findings corroborate previous studies with respect to the persistence of oncogenic infections, while our HPV-level findings do not.

The limitations of this study relate to infection latency, HPV DNA deposition, interval censoring, and estimation of the mean time to detection. First, HPV latency muddles assumptions defined in our analytical frameworks. We attempted to maximize infections acquired during current sexual relationships in analyses of incident infections by including only women/HPV types that were negative at baseline. However, some “incident” infections may be reactivations of latent infections [8]. Latent reactivations have the same HPV positivity signature as incident detections. While the two phenomena are impossible to distinguish, we previously estimated that in the HITCH study, up to 39% of incident infection detections are attributable to latent reactivation [35]. The transition of an infection into a latent state also has the same positivity signature as liberal clearance. To counter the effects of latency, we performed more stringent analyses that reject single-visit reactivation (i.e., double detection) and single-visit latency (i.e., conservative clearance) as events. We attempted to maximize infections acquired during previous relationships in analyses of infections present at baseline by including only women/HPV types that were positive at baseline. However, these infections may have been transmitted by the current male partner in the (up to) 6 months of sexual activity allowed before baseline. Secondly, although women were asked to refrain from sex for 24 hours before providing specimens, a substantial proportion of samples may be false positives as a result of HPV DNA deposition from a male partner [36]. Thirdly, due to interval censoring, discrete times assigned to all HPV results surpass true viral infection and clearance. Our analyses overestimate the times at which incident infections are acquired and present-at-baseline infections are cleared. The time elapsing between detection and clearance of incident infections could be over- or under-estimated, depending on the extent of pre-detection left censoring and post-clearance right censoring. Interval censoring may have also bypassed transient infections (decreasing detection rates) and overlooked clearances that occurred before re-infection (increasing average time to clearance). Fourthly, according to our sensitivity analyses, actuarial mean times to detection were unreliable estimates as a result of heavy right censoring [37]. We were limited in our ability to estimate central tendency for time to detection, since many detection analyses did not reach the 50^th^ percentile required to estimate a median. Fortunately, the actuarial mean was a reliable indicator of average time to clearance.

Our woman-level analyses largely corroborated similar past studies. However, our HPV-level analyses did not clearly indicate that high oncogenic risk subgenus 2 HPV infections are more persistent than their low oncogenic risk and commensal subgenera 1 and 3 counterparts. Our HPV-level estimates of infection natural history provide biologically informed parameters for cervical cancer prevention planning, specifically with respect to the persistence of oncogenic subgenus 2 infections.

## Supporting information

STROBE Cohort

## Data Availability

HITCH participant consent forms specified that data would be published in aggregate form and that individual-level data would only be available to study investigators. To access individual-level data, please contact Eduardo Franco at. Analytical codes and a data dictionary are available on the McGill University Dataverse.

https://doi.org/10.5683/SP3/GVR332

## Acknowledgements

We thank the volunteering participants and employees of the HITCH cohort study, as well as the staff of the Student Health Services Clinics at McGill and Concordia universities. We also thank Cassandra Laurie, Samantha Morais, Suganthiny Jeyaganth, and Talía Malagón for their guidance in selecting and operationalizing statistical analyses. A.W.A. thanks Dr. Vera Hirsh for her contributions to his stipend on behalf of the Hirsh & Schulman Education Fund.

## Additional collaborators in the HITCH study

Affiliated with the Division of Cancer Epidemiology, McGill University, Montréal, Canada: Allita Rodrigues (Study Coordinator); Gail Kelsall, Suzanne Dumais, Natalia Morykon, and Amelia Rocamora (management of subject participation and specimen collection); Nathalie Slavtcheva (study management); Veronika Moravan and Michel Wissing (data management). Affiliated with the Département de Microbiologie Médicale et Infectiologie, Centre Hospitalier de l’Université de Montréal, Montréal, Québec, Canada : Michel Roger (collaborator).

## Author contributions

E.L.F. and A.N.B. designed HITCH and obtained funding. E.L.F. was the principal investigator for the study. A.N.B. oversaw recruitment, data collection, provision of HPV test results to participants, and database design. P.P.T. oversaw clinical activities and recruitment. F.C. supervised laboratory analyses and the quality of polymerase chain reaction assays. M.E.Z. and A.N.B. managed the HITCH database. A.W.A. performed statistical analyses and drafted the manuscript under the supervision of M.E.Z. and E.L.F.. A.N.B. provided statistical analysis consultation. All authors read, provided feedback, and approved the final manuscript.

## Data availability

HITCH participant consent forms specified that data would be published in aggregate form and that individual-level data would only be available to study investigators. To access individual-level data, please contact Eduardo Franco at. The protocol for the HITCH cohort study has been published [24]. Analytical codes and a data dictionary are available on the [McGill University Dataverse](https://borealisdata.ca/dataset.xhtml?persistentId=doi.10.5683/SP3/GVR332).

## Disclaimer

The funders played no role in study design, data collection/analysis, preparation of the manuscript, or the decision to submit it for publication.

## Ethics approval

All subjects provided written informed consent for study participation and use of their biological specimens in future studies. HITCH complies with all national/international regulations regarding research with human data and materials, including the Declaration of Helsinki. The study was conducted per the principles and articles stipulated by the Tri-Council Policy Statement: Ethical Conduct for Research Involving Humans. Ethical approval was obtained from the institutional review boards at McGill University, Concordia University, and the Centre Hospitalier de l’Université de Montréal. Ethics renewal approval is requested annually from McGill University (study number A09-M77-04A).

## Meetings

Portions of this manuscript were presented at a McGill University, Department of Oncology seminar, Montréal, QC, Canada; the 22^nd^ Annual McGill Biomedical Graduate Conference, Montréal, QC, Canada; and the 2022 McGill University, Faculty of Medicine and Health Sciences Celebration of Research and Training in Oncology, Montréal, QC, Canada.

