## Supplementary Material for "Detection and clearance of type-specific and phylogenetically related genital human papillomavirus infections in young women in new heterosexual relationships"

### SUPPLEMENTARY TABLES

**Table S1.** Double detection of incident infection for individual HPV types, grouped types at the woman-level, and grouped types at the HPV-level, by subgenus.

| | n[a] | Cumulative Detection of Infection, % (95% CI) | | | Detection Rate[b] (95% CI) | Time (months) to Detection (95% CI) | | | |
|---|---|---|---|---|---|---|---|---|---|
| | | 6 Months | 12 Months | 24 Months | | Actuarial Mean[c] | Actuarial Median[c] | Conditional Mean[d] | Conditional Median[d] |
| **Subgenus 1** | | | | | | | | | |
| HPV6 | 435 | 0.2 (0.0, 1.7) | 2.0 (1.0, 3.9) | 5.2 (3.4, 8.1) | 2.0 (1.3, 3.0) | 47.4 (46.5, 48.2) | NR | 15.2 (12.2, 18.1) | 15.3 (10.6, 18.1) |
| HPV11[e] | 451 | 0.0 | 0.0 | 0.0 | 0.0 | -- | NR | -- | -- |
| HPV40 | 442 | 0.0[f] | 0.7 (0.2, 2.3) | 2.0 (0.9, 4.1) | 0.6 (0.3, 1.3) | 48.8 (48.3, 49.3) | NR | 14.3 (9.7, 18.9) | 13.1 (6.7, 22.9) |
| HPV42 | 419 | 1.7 (0.8, 3.5) | 4.0 (2.5, 6.5) | 5.9 (3.9, 8.9) | 2.3 (1.5, 3.4) | 47.0 (46.1, 48.0) | NR | 10.8 (7.9, 13.7) | 9.4 (5.4, 11.7) |
| HPV44 | 445 | 0.2 (0.0, 1.6) | 0.5 (0.1, 1.9) | 0.7 (0.2, 2.3) | 0.3 (0.1, 0.8) | 49.1 (48.8, 49.5) | NR | 9.0 (3.8, 14.3) | 7.6 (4.2, NR)[g] |
| HPV54 | 426 | 0.0[f] | 0.8 (0.3, 2.3) | 3.2 (1.8, 5.7) | 1.3 (0.8, 2.3) | 48.0 (47.2, 48.7) | NR | 18.4 (14.8, 22.0) | 18.2 (9.4, 19.7) |
| Any Woman-Level | 367 | 1.7 (0.8, 3.7) | 6.4 (4.2, 9.5) | 13.4 (10.0, 17.7) | 5.9 (4.5, 7.8) | 43.3 (41.8, 44.9) | NR | 15.0 (12.8, 17.2) | 15.3 (9.7, 18.0) |
| Any HPV-Level[h] | 2,618 | 0.3 (0.2, 0.7) | 1.3 (0.9, 1.8) | 2.8 (2.2, 3.7) | 1.1 (0.8, 1.4) | 48.3 (46.3, 48.5) | NR | 14.0 (12.1, 15.9) | 12.6 (9.7, 17.0) |
| **Subgenus 2** | | | | | | | | | |
| HPV16 | 374 | 1.6 (0.7, 3.6) | 2.2 (1.1, 4.4) | 3.9 (2.3, 6.7) | 1.8 (1.1, 2.9) | 47.4 (46.5, 48.4) | NR | 15.1 (10.7, 19.6) | 12.7 (4.8, 21.8) |
| HPV18 | 437 | 0.0[f] | 0.8 (0.2, 2.3) | 1.4 (0.6, 3.3) | 0.6 (0.3, 1.2) | 48.8 (48.3, 49.3) | NR | 15.8 (9.1, 22.5) | 11.4 (8.4, NR)[g] |
| HPV26[e] | 453 | 0.0 | 0.0 | 0.0 | 0.0 | -- | NR | -- | -- |
| HPV31 | 430 | 0.5 (0.1, 1.9) | 1.3 (0.5, 3.0) | 2.4 (1.3, 4.6) | 1.0 (0.6, 1.9) | 48.3 (47.7, 49.0) | NR | 14.5 (10.1, 19.0) | 13.0 (5.1, 19.7) |
| HPV33[e] | 448 | 0.0 | 0.0 | 0.0 | 0.0 | -- | NR | -- | -- |
| HPV34[e] | 450 | 0.0 | 0.0 | 0.0 | 0.0 | -- | NR | -- | -- |
| HPV35 | 450 | 0.2 (0.0, 1.6) | 0.5 (0.1, 1.9) | 0.7 (0.2, 2.2) | 0.3 (0.1, 0.8) | 49.2 (48.8, 49.5) | NR | 8.4 (5.0, 11.8) | 8.5 (4.6, NR)[g] |
| HPV39 | 421 | 0.5 (0.1, 1.9) | 1.0 (0.4, 2.6) | 3.7 (2.1, 6.2) | 1.4 (0.8, 2.3) | 48.0 (47.3, 48.8) | NR | 13.7 (10.5, 16.8) | 12.6 (7.1, 17.2) |
| HPV45 | 445 | 0.0[f] | 0.0[f] | 0.8 (0.3, 2.5) | 0.3 (0.1, 0.8) | 49.2 (48.8, 49.5) | NR | 14.7 (12.8, 16.7) | 14.6 (12.7, NR)[g] |
| HPV51 | 405 | 0.3 (0.0, 1.8) | 1.9 (0.9, 3.9) | 3.4 (1.9, 5.9) | 1.5 (0.9, 2.5) | 47.9 (47.1, 48.6) | NR | 14.9 (11.2, 18.5) | 12.2 (9.3, 18.6) |
| HPV52 | 418 | 0.7 (0.2, 2.2) | 2.0 (1.0, 4.0) | 2.6 (1.4, 4.8) | 1.1 (0.6, 1.9) | 48.3 (47.6, 49.0) | NR | 10.7 (7.1, 14.3) | 9.5 (4.1, 14.0) |
| HPV53 | 419 | 0.5 (0.1, 1.9) | 2.6 (1.4, 4.7) | 4.3 (2.7, 7.0) | 1.8 (1.1, 2.8) | 47.6 (46.7, 48.4) | NR | 12.7 (9.6, 15.7) | 10.6 (8.5, 13.1) |
| HPV56 | 432 | 0.0[f] | 1.3 (0.5, 3.0) | 3.1 (1.7, 5.5) | 1.0 (0.6, 1.9) | 48.4 (47.7, 49.0) | NR | 13.6 (10.7, 16.5) | 13.5 (9.4, 16.3) |
| HPV58 | 431 | 0.2 (0.0, 1.7) | 1.5 (0.7, 3.3) | 2.4 (1.3, 4.6) | 1.0 (0.6, 1.9) | 48.3 (47.6, 49.0) | NR | 14.7 (9.8, 19.7) | 11.7 (7.1, 22.8) |
| HPV59 | 426 | 0.5 (0.1, 1.9) | 0.5 (0.1, 1.9) | 2.2 (1.1, 4.4) | 0.9 (0.5, 1.8) | 48.3 (47.6, 49.0) | NR | 16.9 (11.6, 22.1) | 15.6 (4.8, 21.0) |
| HPV66 | 426 | 1.2 (0.5, 2.8) | 2.2 (1.2, 4.2) | 4.7 (2.9, 7.4) | 1.7 (1.1, 2.7) | 47.6 (46.8, 48.4) | NR | 12.7 (9.2, 16.1) | 11.9 (5.5, 16.9) |
| HPV67 | 427 | 0.5 (0.1, 1.9) | 1.3 (0.5, 3.0) | 3.0 (1.7, 5.3) | 1.1 (0.6, 2.0) | 48.2 (47.5, 48.9) | NR | 13.4 (9.6, 17.3) | 13.1 (5.3, 17.5) |
| HPV68 | 440 | 0.0[f] | 0.7 (0.2, 2.2) | 1.0 (0.4, 2.7) | 0.5 (0.2, 1.1) | 48.9 (48.5, 49.4) | NR | 13.2 (6.2, 20.1) | 7.4 (6.6, NR)[g] |
| HPV69[e] | 453 | 0.0 | 0.0 | 0.0 | 0.0 | -- | NR | -- | -- |
| HPV70 | 450 | 0.0[f] | 0.2 (0.0, 1.7) | 0.5 (0.1, 2.0) | 0.2 (0.0, 0.7) | 49.3 (49.0, 49.5) | NR | 10.6 (6.4, 14.8) | 7.6 (7.6, NR)[g] |
| HPV73 | 435 | 1.2 (0.5, 2.8) | 1.7 (0.8, 3.5) | 3.7 (2.2, 6.2) | 1.4 (0.8, 2.3) | 48.0 (47.2, 48.7) | NR | 12.2 (8.9, 15.5) | 12.3 (4.6, 15.5) |
| HPV82 | 441 | 0.2 (0.0, 1.6) | 0.7 (0.2, 2.2) | 1.0 (0.4, 2.7) | 0.4 (0.1, 1.0) | 49.1 (48.7, 49.4) | NR | 10.8 (4.8, 16.8) | 8.3 (4.5, NR)[g] |
| Any Woman-Level | 233 | 3.9 (2.0, 7.3) | 9.8 (6.6, 14.5) | 21.2 (16.2, 27.5) | 9.5 (7.2, 12.5) | 40.2 (37.9, 42.4) | NR | 14.0 (11.9, 16.1) | 12.8 (10.6, 14.6) |
| Any HPV-Level[h] | 9,511 | 0.4 (0.2, 0.5) | 1.0 (0.7, 1.3) | 2.0 (1.6, 2.5) | 0.8 (0.6, 1.0) | 48.6 (46.6, 48.7) | NR | 13.6 (12.3, 14.8) | 12.3 (11.4, 13.5) |
| **Subgenus 3** | | | | | | | | | |
| HPV61 | 443 | 0.9 (0.4, 2.5) | 1.2 (0.5, 2.8) | 2.7 (1.4, 4.9) | 1.2 (0.7, 2.1) | 48.1 (47.4, 48.8) | NR | 15.7 (10.9, 20.4) | 15.1 (5.4, 22.0) |
| HPV62 | 416 | 0.7 (0.2, 2.3) | 2.3 (1.2, 4.3) | 4.1 (2.5, 6.8) | 1.5 (0.9, 2.4) | 47.9 (47.1, 48.7) | NR | 11.0 (7.9, 14.0) | 9.7 (5.1, 14.7) |
| HPV71[e] | 451 | 0.0 | 0.0 | 0.0 | 0.0 | -- | NR | -- | -- |
| HPV72[e] | 451 | 0.0 | 0.0 | 0.0 | 0.0 | -- | NR | -- | -- |
| HPV81 | 446 | 0.0[f] | 0.2 (0.0, 1.7) | 0.2 (0.0, 1.7) | 0.1 (0.0, 0.6) | 49.3 (49.2, 49.5) | NR | -- | NR |
| HPV83 | 444 | 0.2 (0.0, 1.6) | 0.5 (0.1, 1.9) | 0.5 (0.1, 1.9) | 0.3 (0.1, 0.8) | 49.1 (48.8, 49.5) | NR | 11.7 (1.6, 21.9) | 6.5 (4.3, NR)[g] |
| HPV84 | 413 | 1.5 (0.7, 3.3) | 2.6 (1.4, 4.7) | 4.1 (2.5, 6.7) | 1.7 (1.0, 2.7) | 47.6 (46.8, 48.5) | NR | 12.3 (8.7, 15.9) | 10.6 (5.1, 16.9) |
| HPV89 | 405 | 1.8 (0.8, 3.7) | 3.7 (2.2, 6.1) | 6.8 (4.6, 9.9) | 2.7 (1.9, 4.0) | 46.5 (45.4, 47.6) | NR | 13.2 (10.3, 16.1) | 11.6 (6.9, 18.2) |

| | | | | | | | | | |
|---|---|---|---|---|---|---|---|---|---|
| Any Woman-Level | 336 | 4.2 (2.5, 7.1) | 8.0 (5.5, 11.6) | 13.5 (10.1, 17.9) | 5.8 (4.3, 7.7) | 43.5 (41.9, 45.1) | NR | 12.1 (9.9, 14.4) | 10.1 (6.2, 13.1) |
| Any HPV-Level[h] | 3,469 | 0.6 (0.4, 0.9) | 1.2 (0.9, 1.6) | 2.2 (1.7, 2.8) | 0.9 (0.7, 1.1) | 48.5 (46.5, 48.7) | NR | 12.8 (10.8, 14.9) | 11.4 (7.6, 14.7) |
| **All 36 Types** | | | | | | | | | |
| Any Woman-Level | 192 | 3.7 (1.8, 7.5) | 11.8 (8.0, 17.4) | 30.4 (23.9, 38.1) | 13.3 (10.2, 17.3) | 37.2 (34.6, 39.9) | NR | 14.1 (12.3, 15.9) | 13.1 (12.0, 14.8) |
| Any HPV-Level[h] | 15,598 | 0.4 (0.3, 0.5) | 1.1 (0.9, 1.4) | 2.2 (1.8, 2.6) | 0.9 (0.7, 1.0) | 48.5 (46.5, 48.7) | NR | 13.5 (12.4, 14.5) | 12.2 (10.6, 13.6) |

Abbreviations: CI, confidence interval; HPV, human papillomavirus; NR, not reached.

-- indicates an insufficient number of women were at risk (i.e., 0) or experienced the outcome (i.e., 0 or 1) to estimate this value.

[a] Sample size does not apply to conditional mean/median; sample size for conditional statistics is the number of double detections.

[b] Rate per 1000 woman-months, excepting HPV-level analyses of grouped types, which are provided as rate per 1000 infection-months.

[c] Time to detection including women/types that were censored. Actuarial means were found to be unreliable estimates of average time to detection due to right-censoring.

[d] Time to detection conditional on event of interest (i.e., time to detection restricted to women/types that had a single detection during the study).

[e] HPV type was never detected among women included in this analysis.

[f] HPV type was not detected by this time point among women included in this analysis.

[g] One bound of the survival function's 95% CI never reached or fell below 50%.

[h] 95% CIs for cumulative detection estimates generated using bootstrap resampling of woman-clusters. 95% CIs for detection rate per 1000 infection-months estimated via woman-clustered jackknife. 95% CIs for actuarial and conditional mean time to detection determined by woman-cluster resampling bootstrap. 95% CIs for actuarial and conditional median time to detection affixed by the times at which each (woman-clustered bootstrap-based) 95% CI bound of the actuarial and conditional survival functions (respectively) reached or fell below 50%.

**Table S2.** Conservative clearance of HPV infection present at baseline for individual HPV types, grouped types at the woman-level, and grouped types at the HPV-level, by subgenus.

| | n[a] | Percent of Infection Uncleared, % (95% CI) | | | Clearance Rate[b] (95% CI) | Time (months) to Clearance (95% CI) | | | |
|---|---|---|---|---|---|---|---|---|---|
| | | 6 Months | 12 Months | 24 Months | | Actuarial Mean[c] | Actuarial Median[c] | Conditional Mean[d] | Conditional Median[d] |
| **Subgenus 1** | | | | | | | | | |
| HPV6 | 18 | 69.8 (41.8, 86.3) | 56.4 (29.4, 76.6) | 49.4 (23.5, 70.9) | 31.1 (15.6, 62.2) | 19.9 (13.3, 26.6) | 12.6 (5.7, NR)[e] | 6.6 (4.0, 9.1) | 5.7 (1.4, 11.3) |
| HPV11 | 2 | NR | NR | NR | 228.9 (57.2, 915.0) | 4.4 (2.2, 6.5) | 2.8 (2.8, NR)[e] | 4.4 (2.2, 6.5) | 2.8 (2.8, NR)[e] |
| HPV40 | 11 | 80.8 (42.4, 94.9) | 57.7 (22.1, 81.9) | 19.2 (1.0, 55.4) | 52.5 (23.6, 116.9) | 14.0 (8.7, 19.2) | 12.6 (5.1, NR)[e] | 9.9 (5.3, 14.5) | 7.8 (4.7, NR)[e] |
| HPV42 | 34 | 84.9 (67.4, 93.4) | 50.6 (31.6, 66.9) | 24.3 (9.7, 42.4) | 50.9 (33.2, 78.1) | 16.0 (11.8, 20.1) | 12.2 (8.0, 19.7) | 9.5 (7.7, 11.4) | 8.0 (7.1, 11.7) |
| HPV44 | 8 | 60.0 (19.6, 85.2) | 60.0 (19.6, 85.2) | 45.0 (10.8, 75.1) | 33.4 (12.5, 89.0) | 20.3 (9.6, 30.9) | 13.5 (4.4, NR)[e] | 7.1 (3.5, 10.8) | 4.8 (4.4, NR)[e] |
| HPV54 | 27 | 88.9 (69.4, 96.3) | 58.1 (37.0, 74.3) | 28.4 (11.6, 48.0) | 50.0 (31.9, 78.4) | 16.8 (12.8, 20.7) | 12.5 (8.8, 22.4) | 12.5 (9.3, 15.7) | 10.6 (8.1, 13.7) |
| All Woman-Level | 86 | 80.8 (70.5, 87.8) | 62.5 (50.8, 72.1) | 46.7 (34.6, 57.8) | 33.3 (24.7, 45.0) | 21.1 (18.0, 24.1) | 20.0 (12.2, NR)[e] | 10.0 (8.1, 12.0) | 8.0 (5.8, 11.2) |
| Any HPV-Level[f] | 100 | 79.2 (71.3, 86.4) | 53.9 (40.9, 62.5) | 31.0 (21.6, 42.1) | 46.4 (36.5, 59.3) | 17.5 (14.7, 20.0) | 12.5 (10.6, 14.5) | 9.8 (8.4, 11.2) | 8.0 (7.4, 9.9) |
| **Subgenus 2** | | | | | | | | | |
| HPV16 | 79 | 92.1 (83.3, 96.4) | 79.2 (67.9, 87.0) | 50.9 (37.9, 62.5) | 25.0 (17.9, 35.0) | 24.0 (21.0, 26.9) | 24.0 (18.1, NR)[e] | 12.8 (10.8, 14.8) | 13.6 (8.9, 15.6) |
| HPV18 | 16 | 68.8 (40.5, 85.6) | 50.0 (24.5, 71.1) | 23.4 (6.5, 46.4) | 64.2 (37.3, 110.6) | 13.5 (9.5, 17.5) | 11.9 (5.2, 18.2) | 10.7 (7.3, 14.2) | 11.7 (5.1, 12.4) |
| HPV26 | 0 | -- | -- | -- | -- | -- | -- | -- | -- |
| HPV31 | 23 | 90.9 (68.1, 97.6) | 71.7 (47.6, 86.2) | 34.2 (14.3, 55.4) | 41.6 (24.7, 70.3) | 18.1 (13.5, 22.7) | 13.5 (9.0, 30.2) | 12.1 (8.5, 15.8) | 12.0 (6.2, 13.5) |
| HPV33 | 5 | 60.0 (12.6, 88.2) | NR | NR | 89.4 (28.8, 277.3) | 7.2 (4.7, 9.7) | 7.6 (4.1, NR)[e] | 5.3 (3.6, 7.1) | 4.3 (4.1, NR)[e] |
| HPV34 | 3 | 100.0[g] | 33.3 (0.9, 77.4) | NR | 107.4 (34.6, 333.1) | 9.3 (5.5, 13.1) | 7.3 (6.7, NR)[e] | 9.3 (5.5, 13.1) | 7.3 (6.7, NR)[e] |
| HPV35 | 3 | 100.0[g] | 66.7 (5.4, 94.5) | NR | 43.1 (10.8, 172.5) | 15.7 (8.1, 23.3) | 20.4 (6.2, NR)[e] | 13.3 (3.5, 23.2) | 6.2 (6.2, NR)[e] |
| HPV39 | 32 | 83.2 (64.2, 92.6) | 65.7 (45.5, 79.9) | 22.5 (7.7, 41.9) | 41.6 (26.5, 65.2) | 19.4 (14.2, 24.7) | 17.7 (9.4, 19.7) | 11.9 (9.2, 14.6) | 11.9 (6.0, 17.7) |
| HPV45 | 8 | 75.0 (31.5, 93.1) | 45.0 (10.8, 75.1) | 30.0 (4.4, 62.8) | 52.4 (21.8, 125.8) | 15.3 (7.5, 23.2) | 9.9 (5.5, NR)[e] | 8.4 (6.0, 10.8) | 8.3 (5.5, NR)[e] |
| HPV51 | 48 | 86.7 (72.7, 93.8) | 58.9 (42.2, 72.2) | 26.1 (11.4, 43.5) | 40.1 (27.1, 59.3) | 18.8 (15.0, 22.7) | 17.9 (11.6, 21.8) | 11.2 (8.9, 13.5) | 10.1 (6.1, 13.2) |
| HPV52 | 35 | 87.5 (70.0, 95.1) | 66.0 (45.7, 80.1) | 40.7 (20.6, 60.0) | 32.8 (20.1, 53.6) | 20.2 (15.9, 24.5) | 20.7 (8.7, NR)[e] | 11.9 (8.3, 15.5) | 7.4 (5.1, 17.7) |
| HPV53 | 34 | 93.8 (77.5, 98.4) | 64.4 (44.9, 78.5) | 45.2 (26.4, 62.2) | 30.3 (18.8, 48.7) | 22.5 (17.9, 27.2) | 20.4 (11.5, NR)[e] | 12.7 (9.6, 15.8) | 9.7 (7.8, 17.2) |
| HPV56 | 21 | 85.7 (62.0, 95.2) | 59.2 (34.2, 77.4) | 18.5 (3.4, 43.2) | 45.6 (26.5, 78.5) | 17.6 (11.5, 23.7) | 16.2 (7.8, 21.8) | 10.9 (7.5, 14.2) | 8.1 (5.1, 16.2) |
| HPV58 | 22 | 90.4 (66.8, 97.5) | 62.2 (36.4, 80.0) | 23.3 (6.6, 45.9) | 47.2 (27.4, 81.2) | 16.4 (11.9, 20.8) | 12.9 (8.7, 21.1) | 11.2 (8.6, 13.8) | 9.9 (6.5, 12.9) |
| HPV59 | 27 | 85.2 (65.2, 94.2) | 47.4 (27.0, 65.4) | 12.9 (3.3, 29.4) | 65.0 (42.4, 99.7) | 13.0 (10.4, 15.5) | 11.0 (8.0, 16.2) | 11.0 (8.9, 13.1) | 10.3 (7.7, 14.7) |
| HPV66 | 27 | 73.7 (52.6, 86.5) | 41.0 (22.0, 59.1) | 14.0 (3.7, 31.2) | 73.0 (47.6, 111.9) | 11.6 (8.9, 14.3) | 8.5 (7.5, 14.7) | 9.0 (7.2, 10.8) | 8.0 (4.8, 10.2) |
| HPV67 | 26 | 80.4 (59.2, 91.4) | 51.7 (30.8, 69.1) | NR | 68.3 (44.0, 105.8) | 12.5 (9.9, 15.1) | 12.0 (7.8, 17.7) | 10.7 (8.2, 13.2) | 8.0 (5.4, 14.5) |
| HPV68 | 13 | 100.0[g] | 75.0 (40.8, 91.2) | 50.0 (20.9, 73.6) | 24.3 (10.9, 54.1) | 22.8 (17.3, 28.2) | 19.7 (9.8, NR)[e] | 13.5 (10.4, 16.6) | 11.7 (8.5, NR)[e] |
| HPV69 | 0 | -- | -- | -- | -- | -- | -- | -- | -- |
| HPV70 | 3 | 100.0[g] | 100.0[g] | 50.0 (0.6, 91.0) | 15.6 (2.2, 110.4) | 24.7 (14.6, 34.9) | 17.4 (17.4, NR)[e] | -- | NR |
| HPV73 | 18 | 83.3 (56.8, 94.3) | 43.3 (18.8, 65.7) | 21.6 (5.4, 44.8) | 51.2 (29.1, 90.2) | 15.0 (10.4, 19.6) | 12.0 (9.7, 18.4) | 10.3 (7.7, 12.8) | 9.9 (3.7, 12.6) |
| HPV82 | 12 | 55.0 (23.2, 78.3) | 12.2 (0.7, 40.6) | NR | 116.9 (62.9, 217.2) | 8.0 (5.1, 10.8) | 6.1 (4.1, 11.5) | 7.4 (4.7, 10.1) | 5.5 (2.8, 9.3) |
| All Woman-Level | 220 | 94.8 (90.8, 97.1) | 87.1 (81.6, 91.1) | 64.2 (56.3, 71.0) | 15.6 (12.2, 19.8) | 31.1 (29.1, 33.2) | NR | 14.2 (12.6, 15.9) | 14.6 (12.0, 16.3) |
| Any HPV-Level[f] | 455 | 85.6 (81.6, 88.8) | 60.5 (55.8, 64.8) | 30.1 (24.4, 34.5) | 42.4 (37.8, 47.5) | 20.0 (17.8, 21.4) | 15.5 (13.5, 17.7) | 11.1 (10.4, 11.9) | 9.8 (8.5, 11.0) |
| **Subgenus 3** | | | | | | | | | |
| HPV61 | 10 | 77.8 (36.5, 93.9) | 66.7 (28.2, 87.8) | 53.3 (17.7, 79.6) | 30.7 (12.8, 73.7) | 21.0 (12.5, 29.4) | 26.7 (3.9, NR)[e] | 12.3 (5.0, 19.7) | 9.7 (3.9, NR)[e] |
| HPV62 | 37 | 88.6 (72.4, 95.6) | 67.7 (49.2, 80.7) | 63.5 (44.3, 77.6) | 22.2 (13.1, 37.4) | 24.8 (20.3, 29.4) | NR | 10.7 (6.8, 14.6) | 7.4 (5.9, 8.8) |
| HPV71 | 2 | 100.0[g] | NR | NR | 61.0 (8.6, 433.0) | -- | 9.9 (NR, NR)[e] | -- | NR |
| HPV72[h] | 2 | 100.0 | 100.0 | 100.0 | 0.0 | -- | NR | -- | -- |
| HPV81 | 7 | 100.0[g] | 100.0[g] | NR | 36.2 (11.7, 112.2) | 15.0 (13.8, 16.1) | 14.2 (13.5, NR)[e] | 14.6 (13.3, 15.8) | 14.2 (13.5, NR)[e] |
| HPV83 | 9 | 88.9 (43.3, 98.4) | 77.8 (36.5, 93.9) | 58.3 (15.7, 85.5) | 21.0 (6.8, 65.0) | 24.7 (14.6, 34.9) | NR | 8.1 (4.4, 11.7) | 6.1 (5.5, NR)[e] |
| HPV84 | 40 | 79.4 (63.0, 89.1) | 39.0 (23.1, 54.6) | 25.5 (12.0, 41.4) | 62.3 (43.0, 90.2) | 14.5 (10.5, 18.5) | 8.5 (6.7, 13.1) | 9.2 (6.9, 11.5) | 7.3 (6.1, 8.5) |
| HPV89 | 48 | 87.2 (73.7, 94.0) | 62.4 (46.0, 75.2) | 39.2 (21.7, 56.3) | 35.4 (23.3, 53.7) | 22.3 (17.2, 27.4) | 16.2 (9.7, NR)[e] | 9.6 (7.7, 11.4) | 8.8 (5.7, 11.3) |
| All Woman-Level | 117 | 86.6 (78.7, 91.7) | 71.7 (62.0, 79.3) | 56.4 (45.4, 66.0) | 24.0 (17.9, 32.1) | 26.2 (23.1, 29.3) | NR | 10.3 (8.5, 12.2) | 8.5 (6.1, 10.4) |
| Any HPV-Level[f] | 155 | 85.9 (78.8, 91.0) | 60.2 (49.9, 67.4) | 44.0 (33.7, 52.2) | 35.5 (27.8, 45.4) | 21.8 (18.0, 24.6) | 16.2 (11.3, 25.0) | 10.0 (8.7, 11.3) | 8.2 (6.7, 8.8) |

| | | | | | | | | | |
|---|---|---|---|---|---|---|---|---|---|
| **All 36 Types** | | | | | | | | | |
| All Woman-Level | 261 | 94.9 (91.3, 97.0) | 88.4 (83.6, 91.8) | 74.7 (68.2, 80.1) | 11.8 (9.2, 15.1) | 33.3 (31.5, 35.0) | NR | 13.8 (11.9, 15.7) | 13.1 (9.8, 16.1) |
| Any HPV-Level[f] | 710 | 84.8 (80.7, 87.4) | 59.5 (55.0, 63.3) | 32.9 (28.4, 37.6) | 41.4 (37.1, 46.2) | 20.2 (18.3, 21.5) | 15.3 (13.1, 16.6) | 10.7 (10.1, 11.4) | 8.9 (8.3, 9.9) |

Abbreviations: CI, confidence interval; HPV, human papillomavirus; NR, not reached.

-- indicates an insufficient number of women were at risk (i.e., 0) or experienced the outcome (i.e., 0 or 1) to estimate this value.

[a] Sample size does not apply to conditional mean/median; sample size for conditional statistics is the number of conservative clearances.

[b] Rate per 1000 woman-months, excepting HPV-level analyses of grouped types, which are provided as rate per 1000 infection-months.

[c] Time to clearance including women/infections that were censored.

[d] Time to clearance conditional on event of interest (i.e., time to clearance restricted to women/infections that had a conservative clearance during the study).

[e] Bound(s) of the survival function's 95% CI never reached or fell below 50%.

[f] 95% CIs for uncleared infection estimates generated using bootstrap resampling of woman-clusters. 95% CIs for clearance rate per 1000 infection-months estimated via woman-clustered jackknife. 95% CIs for actuarial and conditional mean time to clearance determined by woman-cluster resampling bootstrap. 95% CIs for actuarial and conditional median time to clearance affixed by the times at which each (woman-clustered bootstrap-based) 95% CI bound of the actuarial and conditional survival functions (respectively) reached or fell below 50%.

[g] HPV type was not cleared by this time point among women included in this analysis.

[h] HPV type was never cleared among women included in this analysis.

**Table S3.** Conservative clearance of incident infection for individual HPV types, grouped types at the woman-level, and grouped types at the HPV-level, by subgenus.

| | n[a] | Percent of Infection Uncleared, % (95% CI) | | | Clearance | Time (months) to Clearance (95% CI) | | | |
|---|---|---|---|---|---|---|---|---|---|
| | | 6 Months | 12 Months | 24 Months | Rate[b] (95% CI) | Actuarial Mean[c] | Actuarial Median[c] | Conditional Mean[d] | Conditional Median[d] |
| **Subgenus 1** | | | | | | | | | |
| HPV6 | 32 | 83.0 (63.9, 92.6) | 47.3 (25.0, 66.7) | 20.3 (1.7, 53.4) | 48.3 (28.6, 81.5) | 14.3 (11.0, 17.7) | 10.5 (8.5, NR)[e] | 8.6 (6.3, 11.0) | 7.6 (4.8, 9.6) |
| HPV11[f] | 1 | NR | NR | NR | 0.0 | -- | NR | -- | -- |
| HPV40 | 14 | 92.3 (56.6, 98.9) | 73.9 (38.5, 90.8) | NR | 24.4 (9.1, 64.9) | 16.7 (13.0, 20.3) | 16.2 (8.7, NR)[e] | 10.0 (6.1, 13.9) | 8.7 (5.2, NR)[e] |
| HPV42 | 34 | 87.3 (69.6, 95.1) | 67.7 (46.6, 81.9) | 41.9 (19.4, 63.0) | 32.1 (18.6, 55.3) | 18.8 (14.8, 22.8) | 16.0 (10.8, NR)[e] | 9.9 (7.5, 12.2) | 8.6 (5.6, 13.7) |
| HPV44 | 4 | 100.0[g] | 66.7 (5.4, 94.5) | NR | 24.7 (3.5, 175.7) | 14.5 (8.6, 20.4) | NR | -- | NR |
| HPV54 | 19 | 89.5 (64.1, 97.3) | 78.3 (42.9, 93.2) | 78.3 (42.9, 93.2) | 14.5 (4.7, 45.0) | 20.8 (16.8, 24.7) | NR | 5.7 (1.9, 9.5) | 3.7 (3.0, NR)[e] |
| All Woman-Level | 69 | 88.1 (77.6, 93.9) | 68.7 (53.9, 79.6) | 41.4 (19.1, 62.6) | 27.8 (18.3, 42.2) | 19.6 (16.5, 22.7) | 22.1 (14.7, NR)[e] | 9.4 (7.4, 11.4) | 8.7 (5.3, 10.8) |
| Any HPV-Level[h] | 104 | 88.4 (81.3, 93.4) | 72.7 (63.0, 79.9) | 64.5 (53.6, 72.1) | 18.3 (13.1, 25.9) | 26.9 (23.5, 29.1) | NR | 8.9 (7.6, 10.5) | NR |
| **Subgenus 2** | | | | | | | | | |
| HPV16 | 22 | 88.5 (61.4, 97.0) | 77.5 (42.1, 92.8) | NR | 26.1 (10.9, 62.7) | 15.2 (11.9, 18.6) | 15.3 (9.0, NR)[e] | 9.5 (5.9, 13.1) | 9.0 (4.8, NR)[e] |
| HPV18 | 10 | 88.9 (43.3, 98.4) | 76.2 (33.2, 93.5) | 76.2 (33.2, 93.5) | 13.9 (3.5, 55.5) | 24.6 (17.8, 31.4) | NR | 6.7 (5.1, 8.3) | 5.6 (5.6, NR)[e] |
| HPV26 | 0 | -- | -- | -- | -- | -- | -- | -- | -- |
| HPV31 | 19 | 94.1 (65.0, 99.2) | 70.6 (38.9, 88.0) | NR | 28.2 (12.7, 62.7) | 15.9 (13.2, 18.6) | 19.7 (8.9, NR)[e] | 11.6 (7.5, 15.6) | 8.9 (5.3, NR)[e] |
| HPV33 | 4 | 50.0 (5.8, 84.5) | NR | NR | 153.1 (57.5, 408.0) | 6.5 (4.5, 8.5) | 5.5 (3.7, NR)[e] | 6.5 (4.5, 8.5) | 5.5 (3.7, NR)[e] |
| HPV34[f] | 2 | 100.0 | NR | NR | 0.0 | -- | NR | -- | -- |
| HPV35 | 4 | 100.0[g] | 100.0[g] | NR | 18.4 (2.6, 130.9) | 20.5 (17.7, 23.3) | 18.5 (18.5, NR)[e] | -- | NR |
| HPV39 | 19 | 94.7 (68.1, 99.2) | 67.6 (37.9, 85.4) | 48.3 (19.5, 72.3) | 30.6 (14.6, 64.2) | 19.6 (14.0, 25.3) | 14.7 (8.5, NR)[e] | 9.1 (6.8, 11.5) | 8.5 (4.7, 12.6) |
| HPV45 | 8 | 87.5 (38.7, 98.1) | 87.5 (38.7, 98.1) | 87.5 (38.7, 98.1) | 10.5 (1.5, 74.5) | 25.5 (20.1, 31.0) | NR | -- | NR |
| HPV51 | 23 | 86.7 (64.3, 95.5) | 70.1 (39.9, 87.2) | 48.1 (17.7, 73.4) | 27.2 (12.9, 57.0) | 19.2 (14.3, 24.1) | 16.3 (11.3, NR)[e] | 9.2 (5.5, 12.9) | 9.4 (3.6, 15.2) |
| HPV52 | 19 | 94.7 (68.1, 99.2) | 53.3 (27.9, 73.3) | 16.7 (1.3, 48.0) | 50.0 (27.7, 90.3) | 14.8 (10.8, 18.7) | 13.7 (6.8, 23.0) | 10.2 (7.0, 13.4) | 9.0 (6.2, 13.7) |
| HPV53 | 25 | 87.5 (66.1, 95.8) | 76.5 (51.8, 89.7) | 68.8 (41.7, 85.2) | 22.2 (10.0, 49.5) | 19.2 (15.5, 22.9) | NR | 6.5 (3.8, 9.1) | 4.2 (3.6, NR)[e] |
| HPV56 | 16 | 85.7 (53.9, 96.2) | 43.5 (14.0, 70.3) | NR | 49.3 (23.5, 103.4) | 11.8 (8.9, 14.7) | 9.2 (7.9, NR)[e] | 8.9 (6.2, 11.5) | 8.2 (5.3, 9.2) |
| HPV58 | 13 | 100.0[g] | 100.0[g] | 66.7 (5.4, 94.5) | 5.2 (0.7, 36.7) | 30.4 (23.9, 36.9) | NR | -- | NR |
| HPV59 | 18 | 82.2 (54.3, 93.9) | 63.4 (31.1, 83.7) | 63.4 (31.1, 83.7) | 27.2 (11.3, 65.4) | 21.8 (15.4, 28.2) | NR | 6.0 (4.2, 7.9) | 5.6 (3.4, NR)[e] |
| HPV66 | 35 | 81.7 (63.7, 91.3) | 47.6 (25.7, 66.6) | NR | 57.8 (35.9, 93.0) | 11.9 (9.5, 14.4) | 10.3 (8.6, 14.9) | 8.7 (6.5, 10.9) | 8.6 (4.6, 10.3) |
| HPV67 | 25 | 100.0[g] | 56.5 (31.2, 75.5) | NR | 41.1 (22.7, 74.2) | 13.3 (11.2, 15.4) | 14.7 (8.3, NR)[e] | 10.2 (8.4, 12.0) | 8.5 (7.4, 13.3) |
| HPV68 | 6 | 80.0 (20.4, 96.9) | 53.3 (6.8, 86.3) | NR | 38.5 (9.6, 153.9) | 12.0 (8.5, 15.4) | NR | 8.0 (4.6, 11.4) | 5.6 (5.6, NR)[e] |
| HPV69 | 0 | -- | -- | -- | -- | -- | -- | -- | -- |
| HPV70[f] | 5 | 100.0 | 100.0 | 100.0 | 0.0 | -- | NR | -- | -- |
| HPV73 | 21 | 95.0 (69.5, 99.3) | 63.3 (35.7, 81.7) | NR | 44.7 (24.1, 83.1) | 14.2 (11.6, 16.8) | 16.3 (8.8, 17.2) | 11.4 (8.7, 14.1) | 9.0 (4.8, 16.3) |
| HPV82 | 9 | 88.9 (43.3, 98.4) | 31.8 (4.9, 64.7) | NR | 58.0 (24.1, 139.3) | 10.6 (7.8, 13.5) | 9.4 (3.8, NR)[e] | 8.0 (6.1, 9.8) | 8.9 (3.8, NR)[e] |
| All Woman-Level | 68 | 95.4 (86.5, 98.5) | 78.6 (64.3, 87.7) | 65.2 (44.2, 80.0) | 16.3 (9.7, 27.5) | 29.8 (25.3, 34.2) | NR | 9.1 (6.5, 11.6) | 8.1 (4.9, 10.1) |
| Any HPV-Level[h] | 303 | 90.4 (86.3, 93.6) | 72.1 (67.1, 77.3) | 61.1 (54.6, 66.8) | 20.5 (16.6, 25.6) | 29.8 (24.6, 31.7) | NR | 9.3 (8.4, 10.2) | NR |
| **Subgenus 3** | | | | | | | | | |
| HPV61 | 16 | 92.9 (59.1, 99.0) | 83.6 (48.0, 95.7) | NR | 17.9 (5.8, 55.4) | 19.0 (14.5, 23.5) | NR | 8.5 (5.1, 11.9) | 7.9 (5.2, NR)[e] |
| HPV62 | 25 | 96.0 (74.8, 99.4) | 78.8 (51.8, 91.7) | 51.7 (22.1, 74.9) | 27.2 (13.6, 54.4) | 20.3 (15.4, 25.2) | 29.0 (12.2, NR)[e] | 12.4 (7.5, 17.3) | 10.8 (3.7, 14.4) |
| HPV71 | 0 | -- | -- | -- | -- | -- | -- | -- | -- |
| HPV72 | 0 | -- | -- | -- | -- | -- | -- | -- | -- |
| HPV81 | 2 | 100.0[g] | 100.0[g] | NR | 44.8 (6.3, 318.2) | -- | 12.2 (NR, NR)[e] | -- | NR |
| HPV83 | 7 | 100.0[g] | 20.0 (0.8, 58.2) | NR | 66.0 (24.8, 175.9) | 9.6 (7.9, 11.3) | 8.8 (7.3, NR)[e] | 8.7 (7.7, 9.8) | 8.5 (7.3, NR)[e] |
| HPV84 | 34 | 79.3 (59.4, 90.2) | 52.6 (31.2, 70.2) | 46.8 (25.4, 65.6) | 39.0 (22.6, 67.1) | 18.1 (13.4, 22.7) | 13.9 (6.9, NR)[e] | 6.9 (5.4, 8.4) | 6.1 (4.5, 8.5) |
| HPV89 | 43 | 90.2 (75.9, 96.2) | 69.5 (51.3, 82.0) | 54.9 (29.0, 74.7) | 25.7 (14.9, 44.3) | 21.0 (17.3, 24.7) | NR | 8.3 (5.9, 10.7) | 6.7 (4.2, 10.5) |
| All Woman-Level | 71 | 89.4 (79.1, 94.8) | 70.6 (56.6, 80.7) | 52.4 (35.2, 67.0) | 29.2 (19.6, 43.6) | 20.3 (17.3, 23.2) | 29.0 (12.9, NR)[e] | 9.4 (7.1, 11.7) | 6.8 (5.7, 10.8) |
| Any HPV-Level[h] | 127 | 90.5 (83.7, 94.5) | 73.0 (64.3, 79.8) | 65.2 (53.8, 72.4) | 19.4 (14.1, 27.1) | 26.6 (23.5, 28.9) | NR | 8.8 (7.3, 10.5) | NR |

| | | | | | | | | | |
|---|---|---|---|---|---|---|---|---|---|
| **All 36 Types** | | | | | | | | | |
| All Woman-Level | 65 | 96.8 (87.7, 99.2) | 85.1 (72.3, 92.3) | 72.1 (50.6, 85.5) | 11.8 (6.5, 21.3) | 31.9 (27.9, 36.0) | NR | 9.7 (6.8, 12.7) | 8.7 (5.3, 12.6) |
| Any HPV-Level[h] | 534 | 90.0 (86.9, 92.4) | 72.5 (68.4, 76.8) | 62.7 (57.9, 66.9) | 19.8 (16.8, 23.4) | 30.1 (25.3, 31.4) | NR | 9.1 (8.4, 9.9) | NR |

Abbreviations: CI, confidence interval; HPV, human papillomavirus; NR, not reached.

-- indicates an insufficient number of women were at risk (i.e., 0) or experienced the outcome (i.e., 0 or 1) to estimate this value.

[a] Sample size does not apply to conditional mean/median; sample size for conditional statistics is the number of conservative clearances.

[b] Rate per 1000 woman-months, excepting HPV-level analyses of grouped types, which are provided as rate per 1000 infection-months.

[c] Time to clearance including women/infections that were censored.

[d] Time to clearance conditional on event of interest (i.e., time to clearance restricted to women/infections that had a conservative clearance during the study).

[e] Bound(s) of the survival function’s 95% CI never reached or fell below 50%.

[f] HPV type was not detected by this time point among women included in this analysis.

[g] HPV type was not cleared by this time point among women included in this analysis.

[h] 95% CIs for uncleared infection estimates generated using bootstrap resampling of woman-clusters. 95% CIs for clearance rate per 1000 infection-months estimated via woman-clustered jackknife. 95% CIs for actuarial and conditional mean time to clearance determined by woman-cluster resampling bootstrap. 95% CIs for actuarial and conditional median time to clearance affixed by the times at which each (woman-clustered bootstrap-based) 95% CI bound of the actuarial and conditional survival functions (respectively) reached or fell below 50%.

**Table S4.** Sensitivity analysis: Actuarial mean time (months) to detection of incident infection with survival functions extended exponentially to 0 for individual HPV types, grouped types at the woman-level, and grouped types at the HPV-level, by subgenus.

| | Single Detection | | Double Detection | |
|---|---|---|---|---|
| | **Mean (95% CI)** | **Successful Reps. (%)** | **Mean (95% CI)** | **Successful Reps. (%)** |
| **Subgenus 1** | | | | |
| HPV6 | 223.1 (117.5, 441.9) | 100.0 | 742.1 (487.6, 1,240.4) | 100.0 |
| HPV11 | 1,844.4 (661.2, 17,133.2) | 97.4 | -- | --- |
| HPV40 | 256.2 (87.7, 958.7) | 100.0 | 2,508.8 (1,336.9, 7,994.4) | 100.0 |
| HPV42 | 193.7 (122.3, 306.4) | 100.0 | 723.5 (475.0, 1,206.4) | 100.0 |
| HPV44 | 914.2 (464.1, 2,932.4) | 100.0 | 6,631.4 (2,814.9, 21,385.5) | 95.5 |
| HPV54 | 377.4 (244.7, 647.1) | 100.0 | 926.4 (530.9, 1,961.8) | 100.0 |
| Any Woman-Level | 101.8 (78.8, 130.7) | 100.0 | 230.4 (171.5, 321.4) | 100.0 |
| Any HPV-Level[a] | 348.9 (236.5, 514.0) | 100.0 | 1,417.2 (1,078.0, 1,916.2) | 100.0 |
| **Subgenus 2** | | | | |
| HPV16 | 290.8 (178.3, 522.6) | 100.0 | 713.4 (427.4, 1,393.3) | 100.0 |
| HPV18 | 827.9 (477.0, 2,206.7) | 100.0 | 2,254.4 (1,087.8, 9,565.9) | 99.5 |
| HPV26 | 18,369.3 (4,677.2, 19,061.6) | 62.6 | -- | -- |
| HPV31 | 276.1 (124.1, 704.0) | 100.0 | 1,426.7 (834.5, 3,639.8) | 100.0 |
| HPV33 | 1,123.2 (388.3, 9,839.7) | 99.7 | -- | -- |
| HPV34 | 8,656.8 (3,300.0, 18,962.7) | 87.0 | -- | -- |
| HPV35 | 2,714.8 (1,203.0, 10,662.2) | 99.8 | 6,815.1 (2,897.6, 21,583.3) | 95.1 |
| HPV39 | 394.6 (224.1, 762.2) | 100.0 | 1,199.3 (748.5, 2,393.7) | 100.0 |
| HPV45 | 286.1 (97.0, 1,638.4) | 100.0 | 6,055.3 (2,603.9, 18,814.3) | 94.3 |
| HPV51 | 194.3 (88.8, 453.3) | 100.0 | 985.3 (606.7, 1,976.5) | 100.0 |
| HPV52 | 192.4 (78.8, 592.6) | 100.0 | 1,620.6 (985.3, 3,669.8) | 100.0 |
| HPV53 | 370.7 (253.7, 560.2) | 100.0 | 883.3 (573.9, 1,567.5) | 100.0 |
| HPV56 | 266.4 (99.5, 1,073.1) | 100.0 | 1,598.5 (961.1, 3,491.8) | 100.0 |
| HPV58 | 843.1 (502.2, 1,695.8) | 100.0 | 1,279.5 (723.0, 3,341.2) | 100.0 |
| HPV59 | 385.4 (187.0, 940.9) | 100.0 | 1,164.2 (585.9, 3,202.9) | 100.0 |
| HPV66 | 158.3 (77.1, 368.3) | 100.0 | 926.6 (608.8, 1,689.3) | 100.0 |
| HPV67 | 419.5 (263.5, 718.4) | 100.0 | 1,302.8 (796.3, 2,736.0) | 100.0 |
| HPV68 | 2,739.6 (1,440.4, 10,049.3) | 99.6 | 3,397.9 (1,647.4, 15,699.2) | 98.9 |
| HPV69 | -- | -- | -- | -- |
| HPV70 | 2,209.5 (1,084.4, 8,069.4) | 99.8 | 9,835.8 (3,851.2, 21,138.3) | 84.7 |
| HPV73 | 370.0 (178.9, 899.8) | 100.0 | 1,188.1 (769.3, 2,254.4) | 100.0 |
| HPV82 | 1,791.4 (1,040.6, 4,279.5) | 100.0 | 4,774.3 (2,205.4, 20,347.1) | 98.2 |
| Any Woman-Level | 46.5 (30.7, 74.7) | 100.0 | 148.0 (106.4, 211.9) | 100.0 |
| Any HPV-Level[a] | 462.6 (305.6, 679.9) | 100.0 | 1,892.1 (1,540.6, 2,339.2) | 100.0 |
| **Subgenus 3** | | | | |
| HPV61 | 562.5 (366.5, 972.5) | 100.0 | 1,160.4 (680.3, 2,448.7) | 100.0 |
| HPV62 | 129.6 (66.0, 459.9) | 100.0 | 1,168.5 (753.2, 2,173.4) | 100.0 |
| HPV71 | -- | -- | -- | -- |
| HPV72 | -- | -- | -- | -- |
| HPV81 | 1,931.4 (888.5, 10,161.0) | 99.6 | 20,643.8 (5,167.0, 21,187.7) | 65.0 |
| HPV83 | 2,608.7 (1,435.7, 9,580.1) | 99.9 | 5,877.5 (2,391.1, 21,533.9) | 93.9 |
| HPV84 | 167.1 (90.7, 313.0) | 100.0 | 918.7 (579.4, 1,639.4) | 100.0 |
| HPV89 | 229.1 (165.8, 325.9) | 100.0 | 530.9 (351.7, 908.0) | 100.0 |
| Any Woman-Level | 56.4 (37.6, 91.8) | 100.0 | 261.2 (189.7, 359.7) | 100.0 |
| Any HPV-Level[a] | 403.9 (281.2, 596.0) | 100.0 | 1,715.7 (1,300.2, 2,258.2) | 100.0 |
| **All 36 Types** | | | | |
| Any Woman-Level | 45.8 (28.9, 71.9) | 100.0 | 106.7 (77.3, 146.1) | 100.0 |
| Any HPV-Level[a] | 426.0 (298.5, 591.6) | 100.0 | 1,753.8 (1,457.7, 2,102.7) | 100.0 |

Abbreviations: CI, confidence interval; HPV, human papillomavirus; reps., replications.
-- indicates an insufficient number of women were at risk (i.e., 0) or experienced the outcome (i.e., 0 or 1) to estimate this value.
[a] 95% CIs for mean times to detection determined by woman-cluster resampling bootstraps.

**Table S5.** Sensitivity analysis: Actuarial mean time (months) to clearance of infection with survival functions extended exponentially to 0 for individual HPV types, grouped types at the woman-level, and grouped types at the HPV-level, by subgenus.

| | Liberal Clearance | | | | Conservative Clearance | | | |
|---|---|---|---|---|---|---|---|---|
| | Infections Present at Baseline | | Incident Infections | | Infections Present at Baseline | | Incident Infections | |
| | **Mean (95% CI)** | **Successful Reps. (%)** | **Mean (95% CI)** | **Successful Reps. (%)** | **Mean (95% CI)** | **Successful Reps. (%)** | **Mean (95% CI)** | **Successful Reps. (%)** |
| **Subgenus 1** | | | | | | | | |
| HPV6 | 13.7 (9.3, 29.9) | 100.0 | 12.1 (9.1, 17.0) | 100.0 | 43.3 (17.8, 107.5) | 100.0 | 17.5 (11.0, 35.5) | 100.0 |
| HPV11 | 4.4 (2.8, 5.9) | 75.6 | -- | -- | 4.4 (2.8, 5.9) | 75.1 | -- | -- |
| HPV40 | 13.0 (6.8, 27.2) | 100.0 | 11.5 (9.1, 14.2) | 100.0 | 16.8 (9.2, 44.6) | 100.0 | 31.5 (13.0, 173.5) | 98.2 |
| HPV42 | 15.0 (11.6, 21.2) | 100.0 | 19.4 (11.9, 32.2) | 100.0 | 21.8 (12.4, 37.9) | 100.0 | 33.0 (16.4, 66.6) | 100.0 |
| HPV44 | 15.3 (7.0, 28.5) | 100.0 | 11.3 (7.2, 18.2) | 94.9 | 40.6 (9.5, 195.6) | 99.7 | 44.4 (7.2, 81.2) | 63.7 |
| HPV54 | 13.3 (10.0, 19.3) | 100.0 | 15.0 (10.9, 26.9) | 100.0 | 16.8 (12.7, 25.9) | 100.0 | 99.4 (32.6, 470.3) | 96.2 |
| All Woman-Level | 15.6 (13.2, 20.7) | 100.0 | 19.2 (13.3, 29.8) | 100.0 | 34.3 (24.0, 50.2) | 100.0 | 33.4 (18.3, 57.8) | 100.0 |
| Any HPV-Level[a] | 13.9 (11.9, 16.5) | 100.0 | 28.9 (20.7, 38.9) | 100.0 | 23.2 (17.0, 32.3) | 100.0 | 80.6 (57.2, 117.3) | 100.0 |
| **Subgenus 2** | | | | | | | | |
| HPV16 | 28.5 (20.2, 39.7) | 100.0 | 21.2 (10.9, 71.9) | 99.9 | 44.6 (30.8, 66.3) | 100.0 | 24.8 (11.6, 112.6) | 99.5 |
| HPV18 | 12.5 (9.3, 18.2) | 100.0 | 40.4 (11.8, 148.8) | 99.5 | 13.5 (9.9, 20.7) | 100.0 | 109.1 (23.4, 285.6) | 87.0 |
| HPV26 | -- | -- | -- | -- | -- | -- | -- | -- |
| HPV31 | 15.5 (11.5, 21.7) | 100.0 | 11.3 (8.5, 19.9) | 100.0 | 21.2 (14.4, 42.8) | 100.0 | 15.9 (13.2, 79.5) | 100.0 |
| HPV33 | 9.9 (4.9, 38.2) | 97.6 | 6.5 (4.6, 8.6) | 97.8 | 9.9 (4.8, 38.2) | 97.4 | 6.5 (4.6, 8.5) | 97.8 |
| HPV34 | 9.3 (6.7, 14.0) | 100.0 | 5.9 (5.8, 6.0) | 100.0 | 9.3 (6.7, 14.0) | 100.0 | -- | -- |
| HPV35 | 15.7 (6.2, 47.6) | 89.8 | 16.6 (7.7, 22.5) | 98.5 | 15.7 (6.2, 47.6) | 90.6 | 36.8 (18.5, 58.2) | 68.4 |
| HPV39 | 14.2 (11.4, 18.4) | 100.0 | 17.5 (10.0, 35.0) | 100.0 | 25.7 (14.3, 43.9) | 100.0 | 39.6 (14.9, 107.3) | 100.0 |
| HPV45 | 9.4 (6.7, 12.5) | 100.0 | 9.4 (6.2, 14.5) | 100.0 | 23.1 (8.1, 80.6) | 100.0 | 212.4 (25.9, 298.4) | 63.6 |
| HPV51 | 16.0 (13.0, 20.7) | 100.0 | 13.7 (9.8, 17.9) | 100.0 | 25.8 (15.9, 41.6) | 100.0 | 37.6 (15.1, 117.8) | 99.9 |
| HPV52 | 13.9 (10.3, 20.2) | 100.0 | 15.4 (10.0, 27.0) | 100.0 | 28.7 (17.5, 54.4) | 100.0 | 17.2 (10.8, 33.7) | 100.0 |
| HPV53 | 18.5 (15.0, 22.6) | 100.0 | 13.7 (10.7, 23.1) | 100.0 | 34.2 (19.8, 66.3) | 100.0 | 64.4 (26.5, 188.3) | 100.0 |
| HPV56 | 15.8 (9.9, 30.8) | 100.0 | 10.7 (8.2, 15.2) | 100.0 | 22.0 (11.8, 45.8) | 100.0 | 11.8 (9.3, 34.4) | 100.0 |
| HPV58 | 18.3 (11.7, 30.6) | 100.0 | 21.5 (15.4, 29.4) | 99.9 | 21.4 (12.4, 41.0) | 100.0 | 87.1 (22.3, 155.7) | 63.2 |
| HPV59 | 11.5 (9.2, 14.4) | 100.0 | 13.7 (8.1, 21.3) | 100.0 | 14.6 (10.7, 20.8) | 100.0 | 64.4 (16.3, 231.0) | 99.6 |
| HPV66 | 10.9 (8.7, 13.3) | 100.0 | 9.7 (8.0, 11.6) | 100.0 | 13.4 (9.4, 20.8) | 100.0 | 11.9 (9.6, 17.5) | 100.0 |
| HPV67 | 10.7 (8.6, 13.3) | 100.0 | 10.9 (9.1, 13.6) | 100.0 | 13.1 (10.1, 17.6) | 100.0 | 18.3 (12.0, 35.3) | 100.0 |
| HPV68 | 29.6 (15.6, 66.3) | 100.0 | 14.0 (7.9, 52.1) | 98.0 | 45.9 (21.3, 133.2) | 99.8 | 24.8 (8.5, 82.6) | 91.4 |
| HPV69 | -- | -- | -- | -- | -- | -- | -- | -- |
| HPV70 | 18.0 (4.7, 32.0) | 95.2 | 14.4 (6.2, 23.2) | 99.7 | 47.8 (17.4, 111.9) | 65.7 | -- | -- |
| HPV73 | 11.3 (7.7, 16.9) | 100.0 | 11.0 (8.8, 13.4) | 100.0 | 19.4 (10.9, 35.4) | 100.0 | 15.6 (11.8, 26.5) | 100.0 |
| HPV82 | 7.1 (5.1, 9.6) | 100.0 | 11.9 (7.2, 23.7) | 100.0 | 8.0 (5.4, 11.4) | 100.0 | 15.0 (8.0, 41.1) | 99.5 |
| All Woman-Level | 30.9 (23.9, 41.4) | 100.0 | 21.5 (13.6, 33.4) | 100.0 | 77.0 (57.0, 98.9) | 100.0 | 90.0 (39.1, 185.4) | 100.0 |
| Any HPV-Level[a] | 15.5 (14.1, 17.8) | 100.0 | 25.4 (18.2, 31.9) | 100.0 | 27.9 (23.5, 32.7) | 100.0 | 82.5 (61.0, 103.3) | 100.0 |
| **Subgenus 3** | | | | | | | | |
| HPV61 | 24.0 (11.4, 73.2) | 100.0 | 23.1 (9.8, 66.7) | 99.9 | 28.2 (12.9, 103.6) | 99.9 | 58.3 (16.0, 289.7) | 96.6 |
| HPV62 | 20.5 (15.8, 28.6) | 100.0 | 13.1 (9.4, 17.0) | 100.0 | 52.0 (27.7, 103.2) | 100.0 | 20.3 (15.3, 62.2) | 100.0 |
| HPV71 | 8.2 (6.5, 9.9) | 100.0 | -- | -- | -- | -- | -- | -- |
| HPV72 | -- | -- | -- | -- | -- | -- | -- | -- |
| HPV81 | 12.6 (8.2, 27.4) | 99.9 | -- | -- | 15.0 (13.6, 39.2) | 98.6 | -- | -- |
| HPV83 | 26.6 (10.9, 85.4) | 100.0 | 10.4 (7.6, 20.8) | 100.0 | 63.7 (13.7, 304.4) | 97.7 | 11.2 (8.1, 32.4) | 99.9 |
| HPV84 | 11.0 (8.2, 14.8) | 100.0 | 11.5 (8.6, 15.5) | 100.0 | 17.2 (11.0, 28.5) | 100.0 | 36.6 (16.7, 71.8) | 100.0 |
| HPV89 | 12.3 (9.9, 15.3) | 100.0 | 14.4 (11.7, 17.3) | 100.0 | 39.1 (18.8, 65.5) | 100.0 | 47.8 (22.0, 107.4) | 100.0 |

| | | | | | | | | |
|---|---|---|---|---|---|---|---|---|
| All Woman-Level | 18.4 (15.8, 23.8) | 100.0 | 15.3 (13.1, 18.5) | 100.0 | 55.7 (38.5, 77.5) | 100.0 | 26.1 (18.1, 65.0) | 100.0 |
| Any HPV-Level[a] | 14.9 (12.6, 18.2) | 100.0 | 22.8 (17.1, 29.6) | 100.0 | 34.7 (24.3, 47.3) | 100.0 | 72.2 (46.9, 109.5) | 100.0 |
| **All 36 Types** | | | | | | | | |
| All Woman-Level | 32.3 (24.9, 43.8) | 100.0 | 41.3 (21.7, 78.4) | 100.0 | 104.1 (77.3, 137.8) | 100.0 | 118.8 (49.4, 269.3) | 100.0 |
| Any HPV-Level[a] | 15.0 (13.7, 16.5) | 100.0 | 26.3 (20.1, 31.6) | 100.0 | 28.6 (24.1, 32.7) | 100.0 | 84.5 (63.7, 99.9) | 100.0 |

Abbreviations: CI, confidence interval; HPV, human papillomavirus; reps., replications.

-- indicates an insufficient number of women were at risk (i.e., 0) or experienced the outcome (i.e., 0 or 1) to estimate this value.

[a] 95% CIs for mean times to clearance determined by woman-cluster resampling bootstraps.

### SUPPLEMENTARY FIGURES

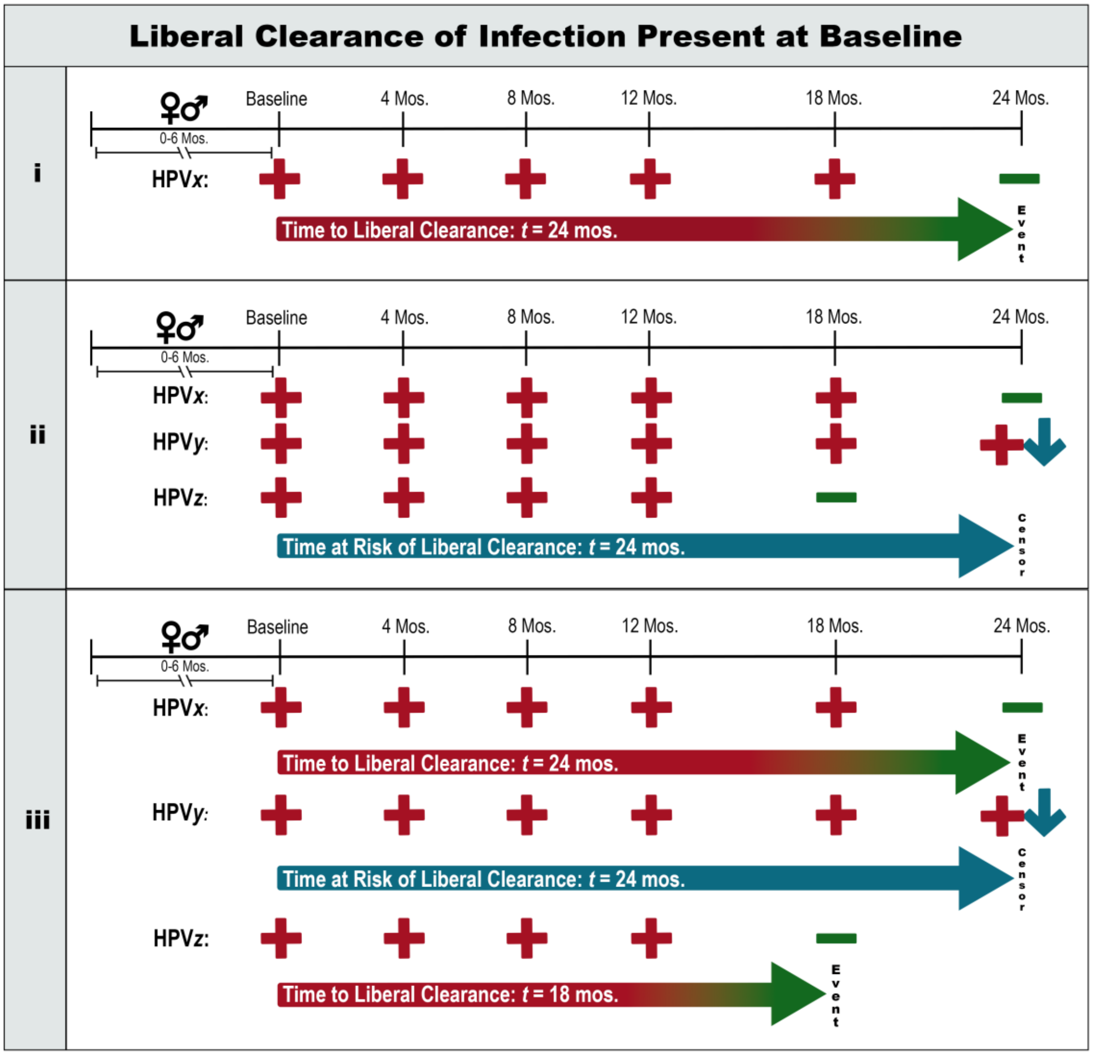


Abbreviations: HPV, human papillomavirus; mos., months (1 month = 30.437 days).

**Figure S1.** Analytical framework for liberal clearance of infection present at baseline.

**i.** Woman-level analysis for a given HPV type: Woman must be positive for HPV*x* at baseline. She has a liberal clearance at 24 months, when she tests negative for HPV*x*.

**ii.** Woman-level analysis for grouped HPV types *x*, *y*, and *z*: Woman must be positive for one or more types at baseline. She never clears all three types at a single visit, so she is right-censored (24 months).

**iii.** HPV-level analysis for grouped HPV types *x*, *y*, and *z*: HPV types must be present at baseline. There will be a liberal clearance each first time woman tests negative for any type following a visit where she was positive for the same type. HPV*x* and HPV*z* have liberal clearances at 24- and 18-months, respectively; HPV*y* is right-censored (24 months).

Symbol Legend:

♀♂ : Debut of woman's sexual relationship with a male partner occurred 0-6 months pre-baseline.

⬇ : Data are right-censored.

⇨ : Time to discrete event of interest. Gradient arrows indicate that biological clearance occurred at an unknown time before the clearance event at the arrowhead. Solid blue arrow counts time at risk contributed before censorship at the arrowhead.

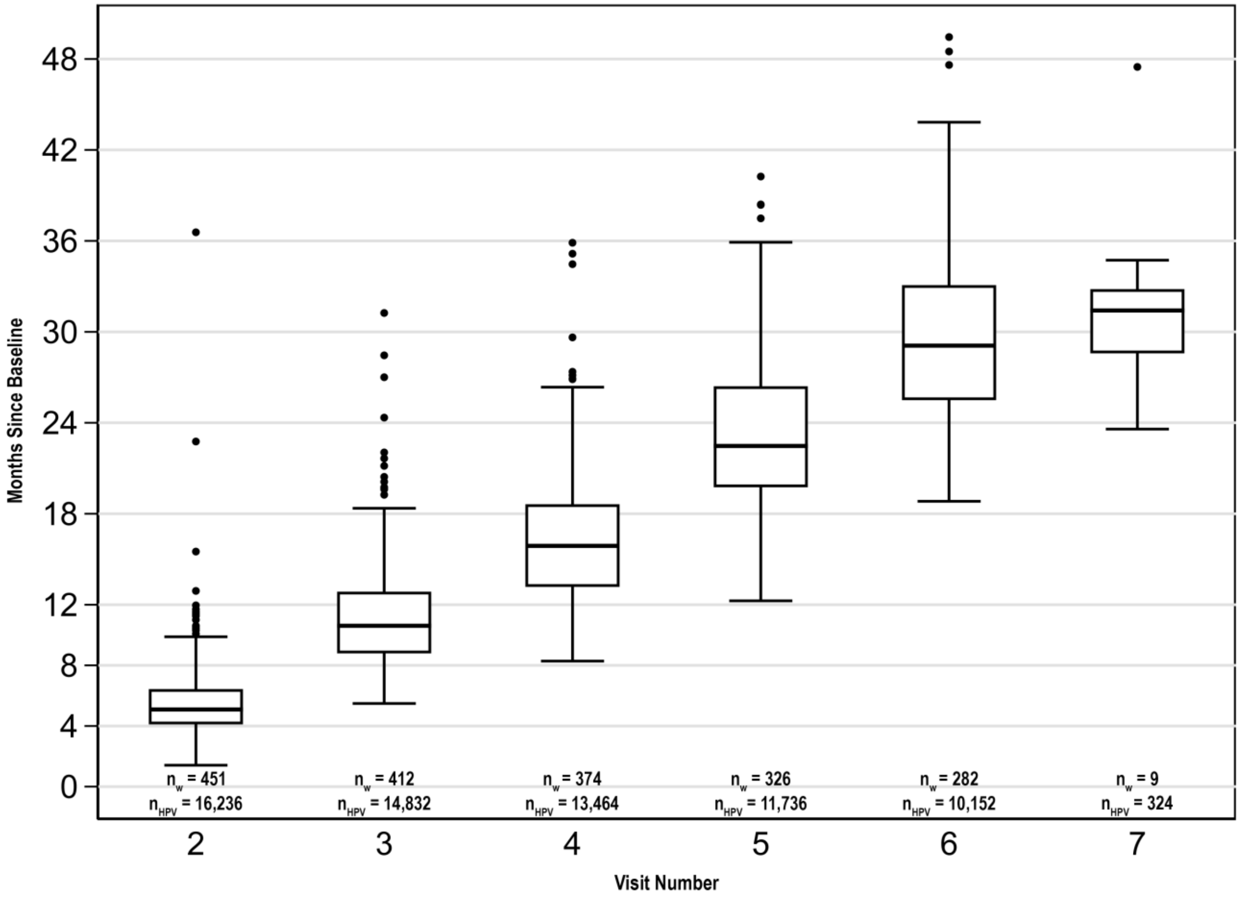


**Figure S2.** Months post-baseline at which women attended follow-up visits. Sample size per visit provided for woman- ($n_w$) and HPV- ($n_{HPV}$) level prevalence analyses. Protocol-designated times for visits 2-6 were 4-, 8-, 12-, 18- and 24-months post-baseline; visit 7 was auxiliary.

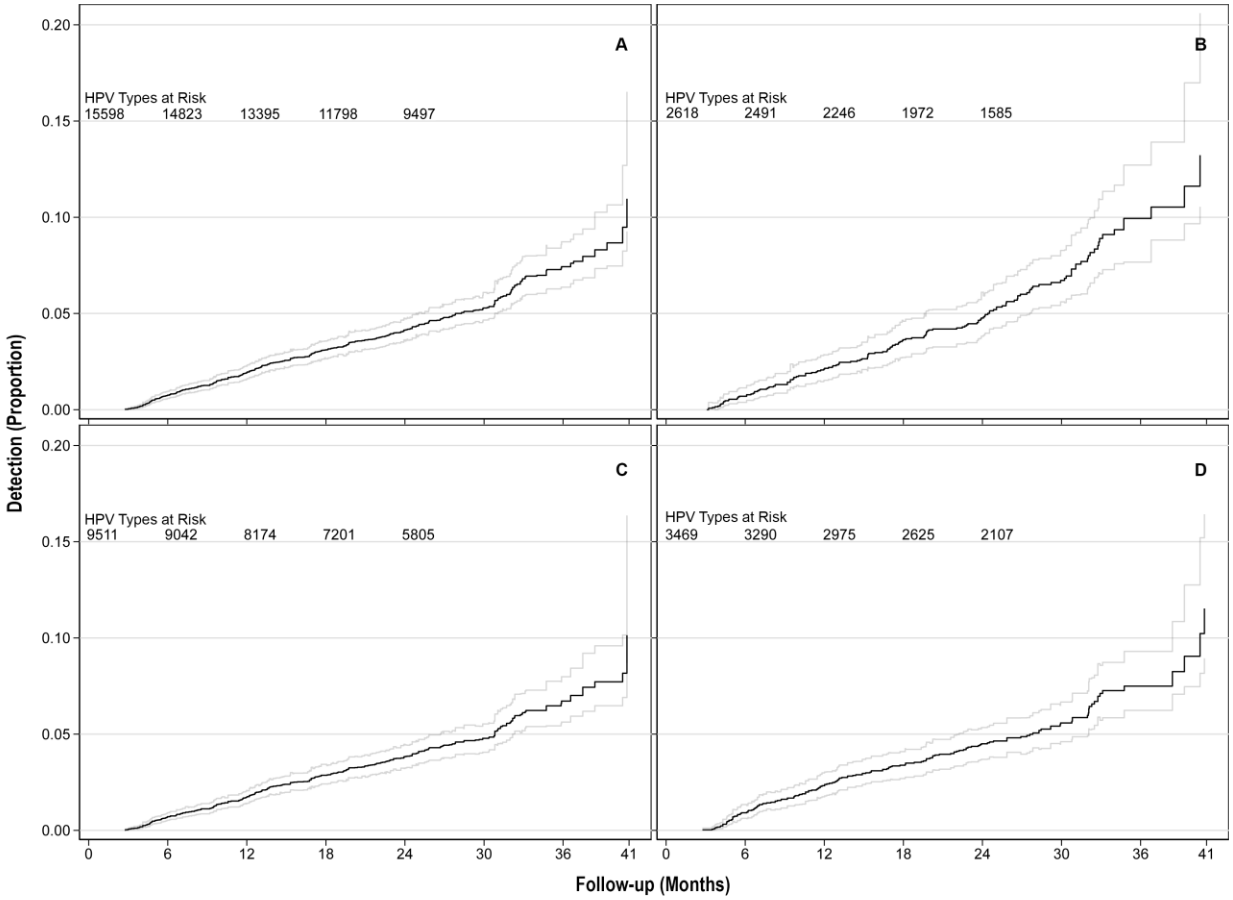


**Figure S3.** Single detection of incident infection with any (A) HPV type, (B) subgenus 1, (C) subgenus 2, and (D) subgenus 3 type, at the HPV-level. Risk tables were extracted from standard Kaplan-Meier plots and appended to the above plots, which incorporate bootstrap-based confidence intervals.

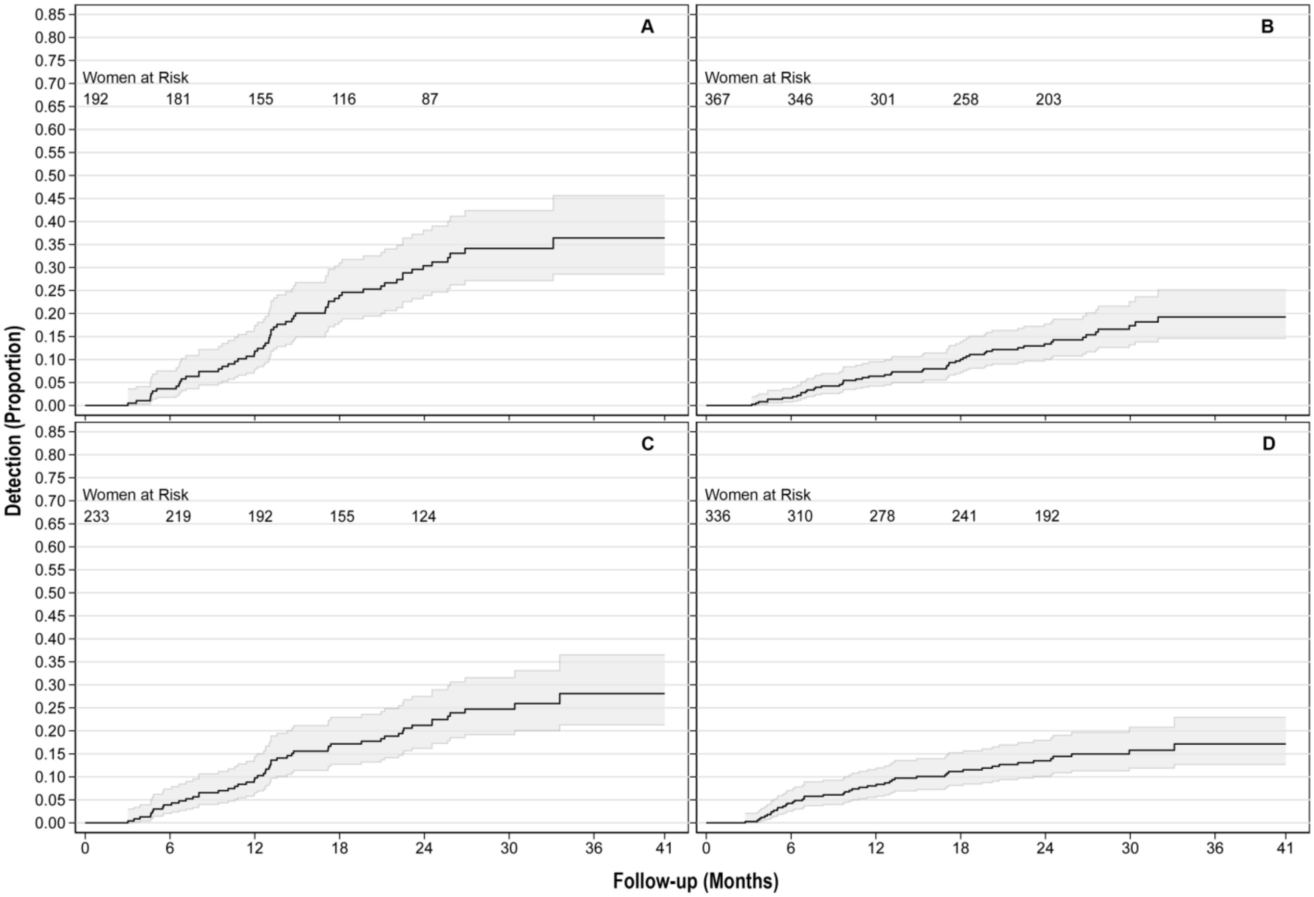


**Figure S4.** Double detection of incident infection with any (A) HPV type(s), (B) subgenus 1, (C) subgenus 2, and (D) subgenus 3 type(s), at the woman-level.

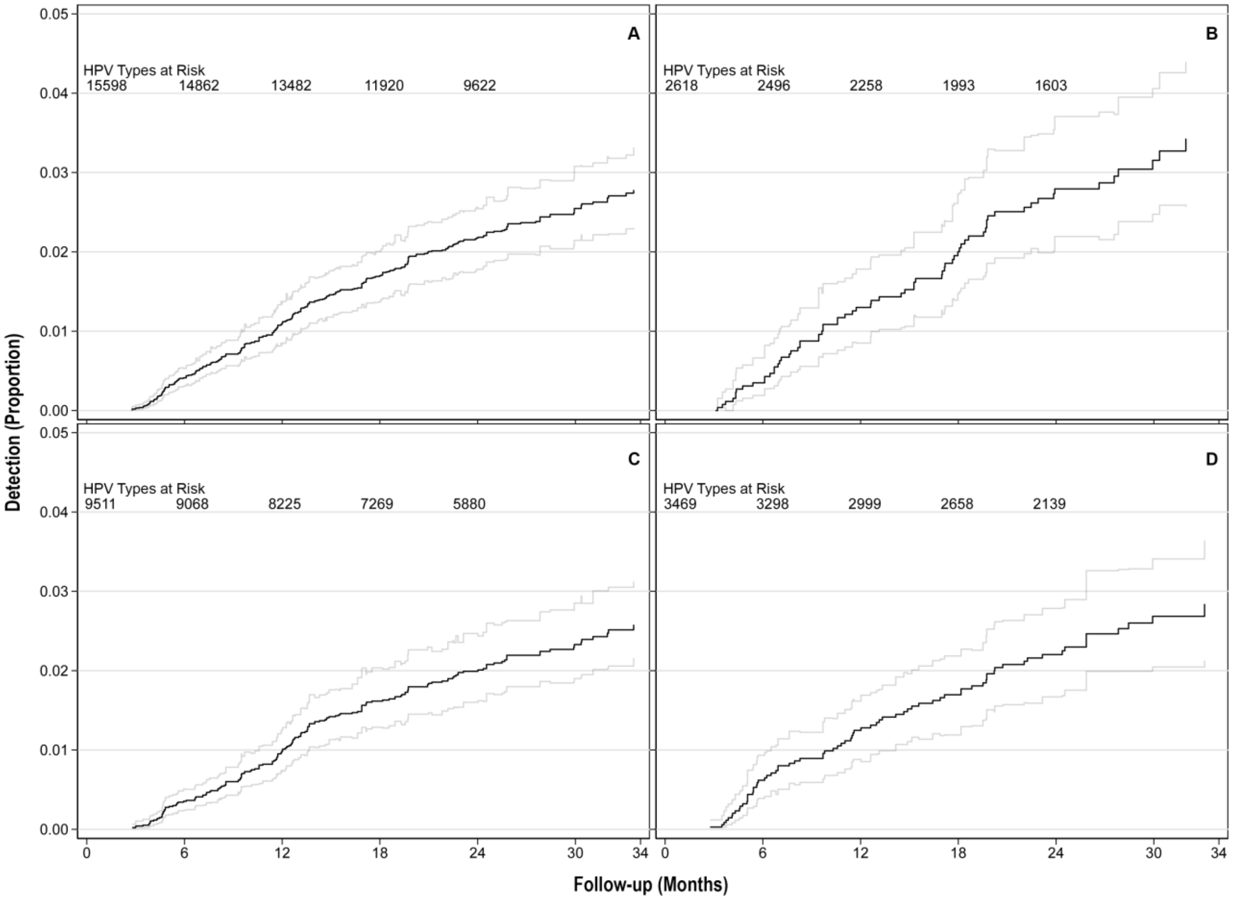


**Figure S5.** Double detection of incident infection with any (A) HPV type, (B) subgenus 1, (C) subgenus 2, and (D) subgenus 3 type, at the HPV-level. Risk tables were extracted from standard Kaplan-Meier plots and appended to the above plots, which incorporate bootstrap-based confidence intervals.

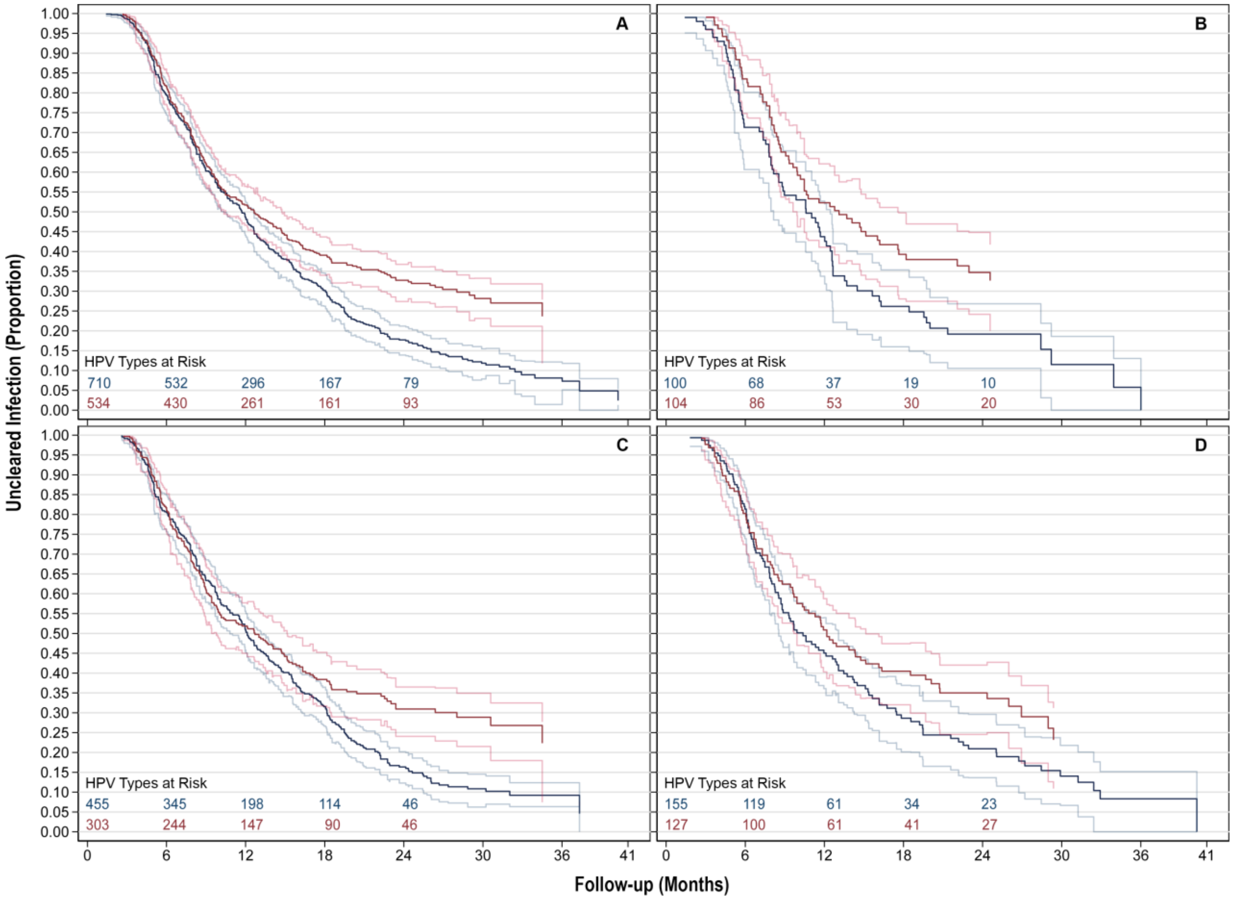


**Figure S6.** Liberal clearance of any (A) HPV type, (B) subgenus 1, (C) subgenus 2, and (D) subgenus 3 type for incident infections (red) and infections present at baseline (blue), at the HPV-level. Risk tables were extracted from standard Kaplan-Meier plots and appended to the above plots, which incorporate bootstrap-based confidence intervals.

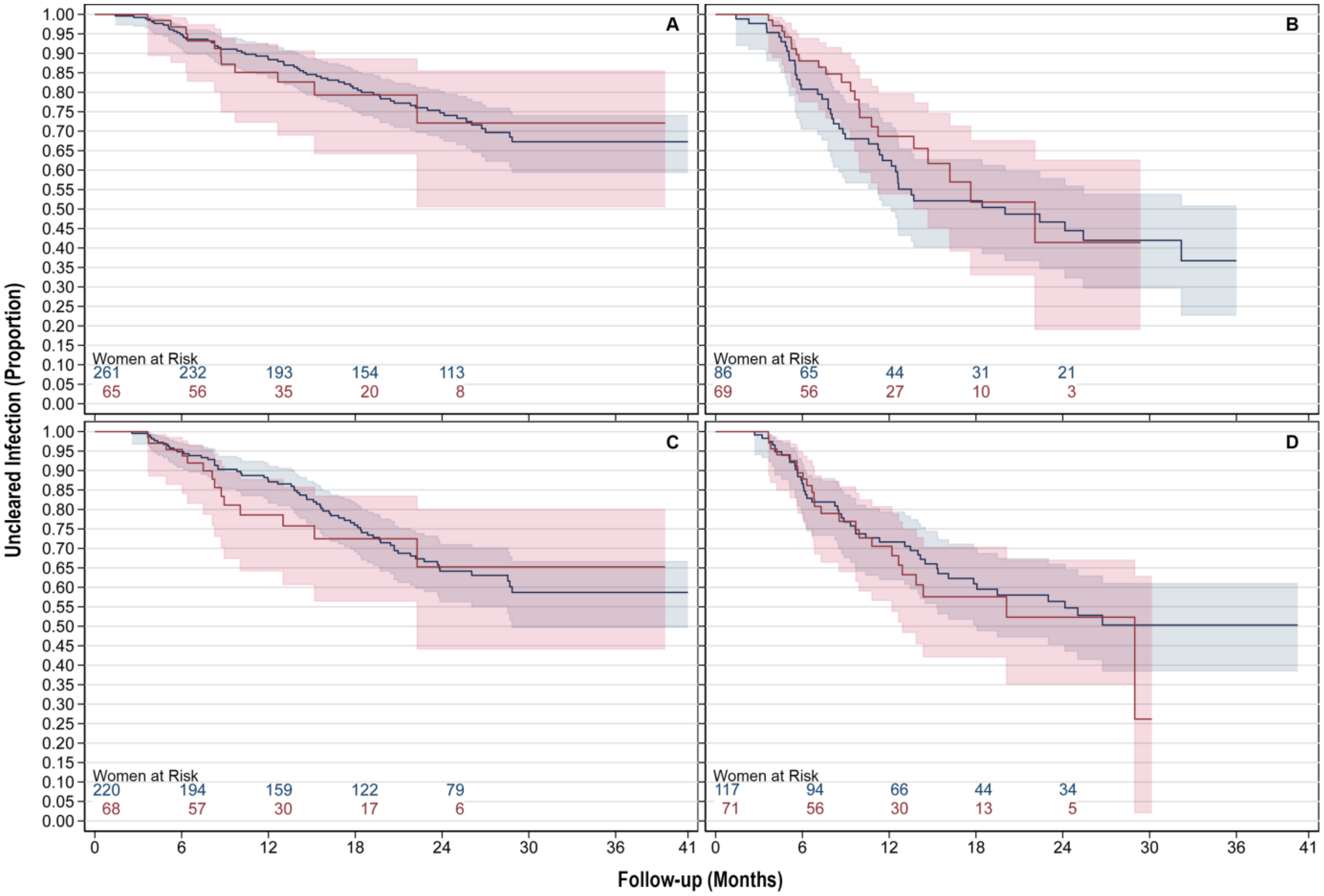


**Figure S7.** Conservative clearance of all (A) HPV type(s), (B) subgenus 1, (C) subgenus 2, and (D) subgenus 3 type(s) for infections present at baseline (blue) and incident infections (red), at the woman-level.

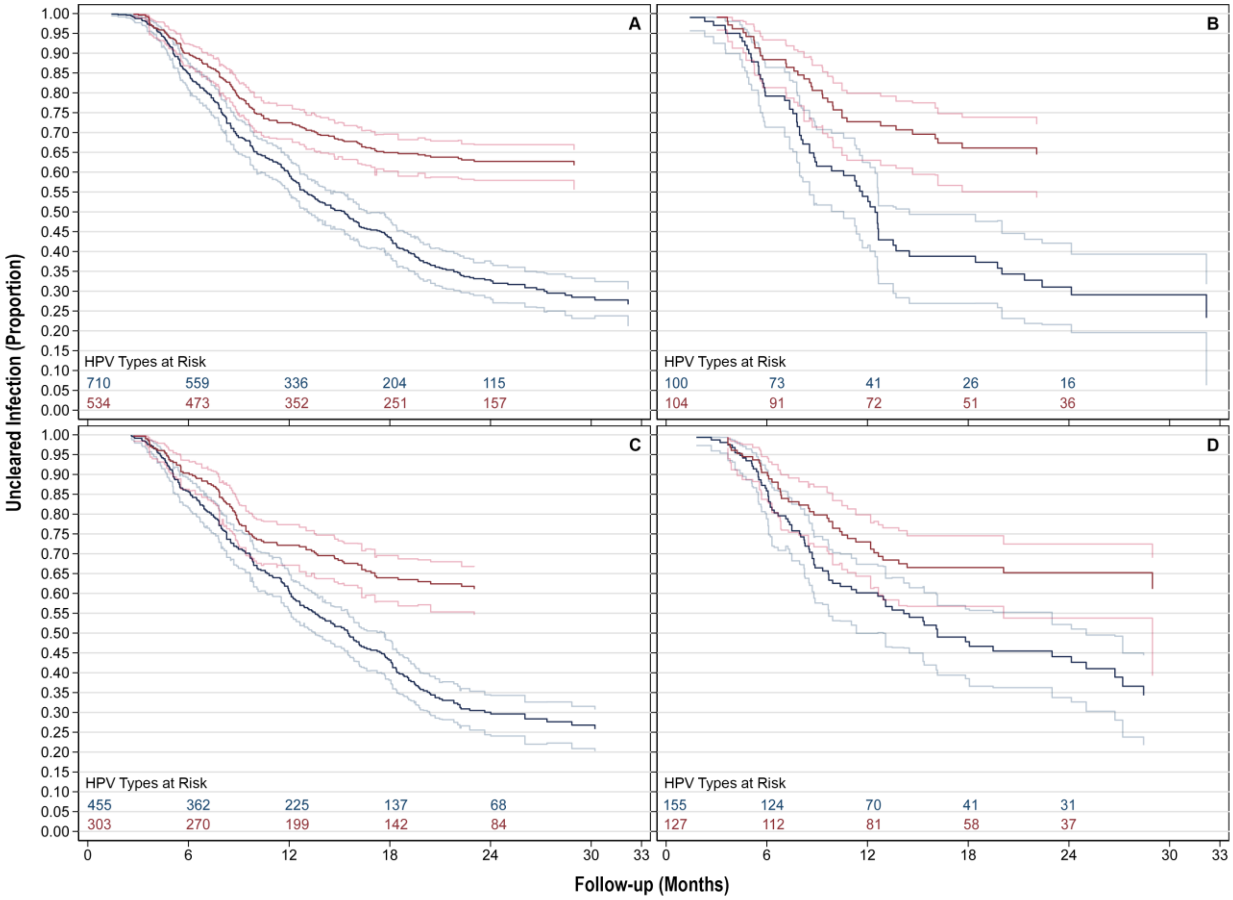


**Figure S8**. Conservative clearance of any (A) HPV type, (B) subgenus 1, (C) subgenus 2, and (D) subgenus 3 type for incident infections (red) and infections present at baseline (blue), at the HPV-level. Risk tables were extracted from standard Kaplan-Meier plots and appended to the above plots, which incorporate bootstrap-based confidence intervals.

**Addenda**

**Addendum S1.** Woman-Level Analyses of Given HPV Types: For each outcome, we performed 36 woman-level analyses of individual HPV types, with one analysis for each HPV type tested. Each analysis of a given HPV type incorporated women's longitudinal positivity statuses for one individual type of HPV.

**Addendum S2.** Woman-Level Analyses of Grouped HPV Types: For each outcome, we performed 4 woman-level analyses of grouped HPV types, with one analysis for each subgenus, and one analysis including all 36 HPV types. These analyses treat each woman's longitudinal HPV status as a composite. A woman was considered HPV-positive when she tested positive for one or more of the HPV types included in the group of interest, and HPV-negative only when she tested negative for *all* those types.

**Addendum S3.** HPV-Level Analyses of Grouped HPV Types: For each outcome, we performed 4 HPV-level analyses of grouped HPV types, with one analysis for each subgenus, and one analysis including all 36 HPV types. These analyses handle women's longitudinal HPV statuses for each unique HPV type in the group of interest separately. In so doing, HPV-level analyses can account for a woman's simultaneous positivity and negativity for multiple different HPV types.
